# When Case Reporting Becomes Untenable: Can Sewer Networks Tell Us Where COVID-19 Transmission Occurs?

**DOI:** 10.1101/2022.09.29.22280508

**Authors:** Yuke Wang, Pengbo Liu, Jamie VanTassell, Stephen P. Hilton, Lizheng Guo, Orlando Sablon, Marlene Wolfe, Lorenzo Freeman, Wayne Rose, Carl Holt, Mikita Browning, Michael Bryan, Lance Waller, Peter F.M. Teunis, Christine L. Moe

## Abstract

Monitoring SARS-CoV-2 in wastewater is a valuable approach to track COVID-19 transmission. Designing wastewater surveillance (WWS) with representative sampling sites and quantifiable results requires knowledge of the sewerage system and virus fate and transport. We developed a multi-level WWS system to track COVID-19 in Atlanta using an adaptive nested sampling strategy. From March 2021 to April 2022, 868 wastewater samples were collected from influent lines to wastewater treatment facilities and upstream community manholes. Variations in SARS-CoV-2 concentrations in influent line samples preceded similar variations in numbers of reported COVID-19 cases in the corresponding catchment areas. Community sites under nested sampling represented mutually-exclusive catchment areas. Community sites with high SARS-CoV-2 detection rates in wastewater covered high COVID-19 incidence areas, and adaptive sampling enabled identification and tracing of COVID-19 hotspots. This study demonstrates how a well-designed WWS provides actionable information including early warning of surges in cases and identification of disease hotspots.

## Introduction

Since the early stages of the COVID-19 pandemic, wastewater surveillance (WWS) has been recognized as a valuable tool that can complement traditional epidemiological surveillance, to monitor the spread of COVID-19 and indicate trends in infection at different levels [1, 2, 3, 4]. Ideally, traditional epidemiological surveillance through monitoring the number of people who tested positive for the SARS-CoV-2, would provide timely and accurate estimation of COVID-19 incidence. However, many factors like asymptomatic infections, non-specific symptoms, diagnostic test sensitivity, limited diagnostic test reagents and testing centers, inadequate data collection and overburdened reporting systems, and public health policies and human behavior could contribute to under-ascertainment, under-reporting, and delayed reporting of COVID-19 using standard disease surveillance approaches [5, 6]. With detection of SARS-CoV-2 RNA in wastewater [7, 8, 9] and in stools from COVID-19 patients [10, 11] during the early COVID-19 pandemic, WWS, previously utilized to monitor enteric diseases [12, 13, 14, 15], was proposed as an inclusive, non-intrusive, inexpensive, sensitive, and scalable strategy to guide public health response to the COVID-19 pandemic [16, 17]. By September 2022, federal and local governments, health departments, municipalities, and universities in at least 70 countries had utilized WWS to monitor COVID-19 [18]. The results of WWS have been used to alert local jurisdictions, guide resource allocation, enable targeted communications, and forecast clinical resource needs [3].

Most WWS studies have monitored and estimated the COVID-19 incidence at the city level [1, 19, 20] by collecting repeated grab or composite wastewater samples from downstream sites (e.g., inlets of wastewater treatment facilities), that serve a large population. When COVID-19 is spreading, wastewater samples from downstream sites consistently contain detectable levels of SARS-CoV-2 RNA. To monitor temporal trends of COVID-19 incidence in the catchment population, quantification of SARS-CoV-2 RNA concentration in wastewater is required. WWS has also been widely used at the institution level to monitor disease transmission and provide early warning of surges in cases or outbreaks [4, 21, 22, 23]. Usually, WWS for early warning is considered when the incidence is low. Frequent sampling is necessary to enable a quick confirmation of an outbreak and a timely response. And the catchments of sampling sites would preferably be small, which enables targeted control measures. However, few WWS studies have focused on sampling upstream sites at the community/neighborhood level [24, 25]. Sampling upstream has the potential to identify disease hotspots within a larger area, which enables monitoring localized disease transmission and targeted public health interventions [25, 26]. Unlike sampling at wastewater treatment facilities, sample collection at community sites across the sewer network is more complicated and requires more strategic sampling design in order to optimize the value of information and guide interpretation of the results [15, 27, 28, 29, 30, 31].

In this study, we conducted COVID-19 WWS at the city and community level in the city of Atlanta. The objectives were to: (1) strategically design the WWS sampling by describing the sewer network in the city of Atlanta from a disease monitoring perspective, and developing and applying an algorithm to select and adapt community sampling sites across sewer networks; (2) examine the association between WWS results and COVID-19 incidence at different levels; and (3) illustrate the capacity of WWS to detect and monitor spatial COVID-19 hotspots at the community level. The knowledge gained in this study demonstrates the value of strategic sampling design for WWS to provide actionable information to guide public health response to COVID-19.

## Results

### Atlanta Sewer Network

We examined the network statistics (indegree and outdegree) and network topology of the Atlanta sewerage system. The indegree and outdegree of a manhole in the sewer network are the numbers of sewer pipes going into and going out of the manhole, respectively. The city of Atlanta sewer network has a total of 52,787 sewer segments and 51,155 manholes. After excluding nodes with zero indegree or zero outdegree, the average indegree and outdegree were 1.40 and 1.04 respectively indicating it was mainly a tree topology network with some net topology structures. However, the sewer network topology varied in different areas. In newly developed areas with low population density, the sewer network includes more line topology components. In densely populated inner city areas with older sewerage structures, there are more cross-connections and greater connectivity. Figure 1 illustrates two sub-networks in our study: one near the center of the city (upstream of manhole 23370330601) and the other located more remotely (upstream of manhole 23330211001). The average indegree and outdegree were larger for the sewer network in the city center, which displays a more connected structure. Figures 2a and 2b show the spatial distribution of in- and outdegree in the Atlanta sewer network. The nodes with large indegree and outdegree are located mainly at the center of the city where the sewers are mainly combined sewers (Figure 2c).

**Figure 1:**
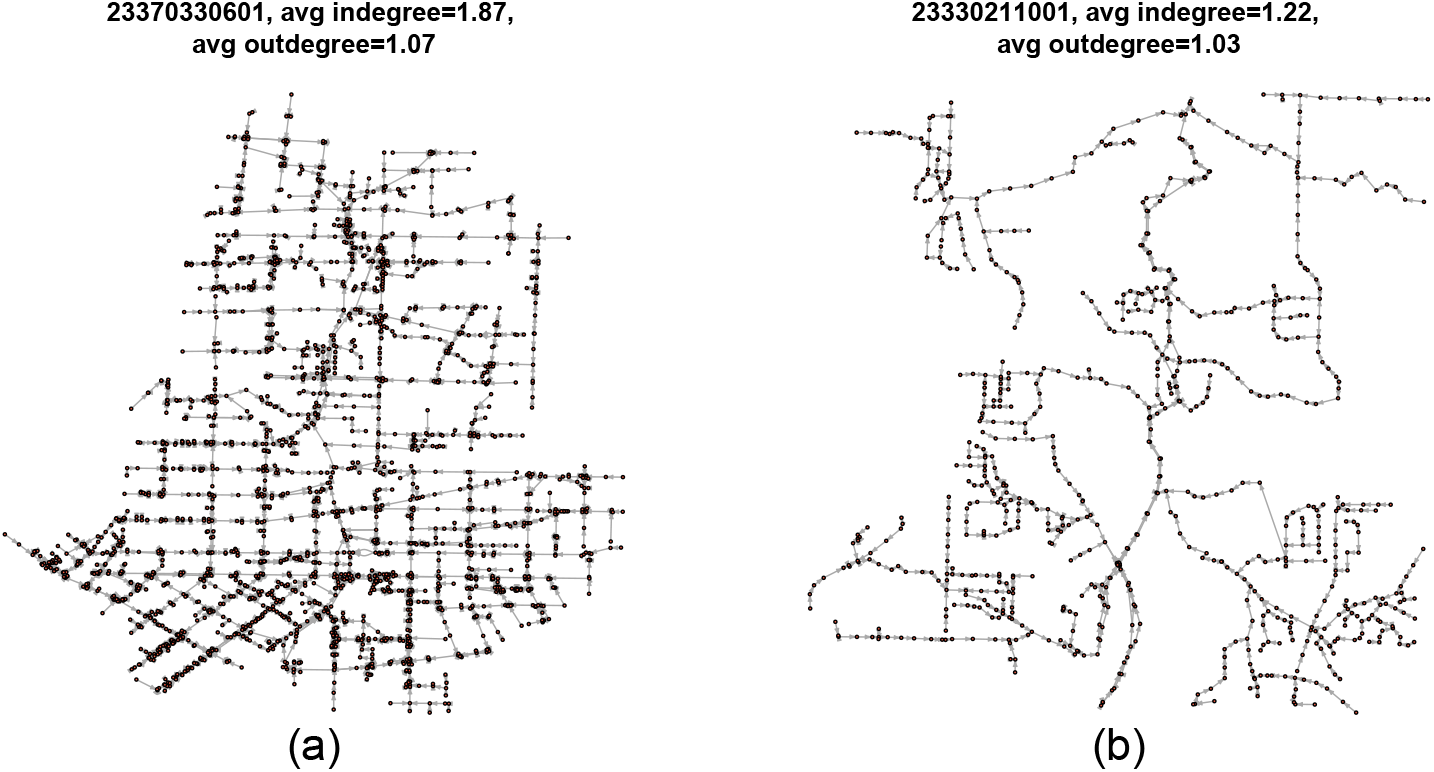
Two sewer sub-networks (a) within the center of the city and (b) in the periphery of the city.

**Figure 2:**
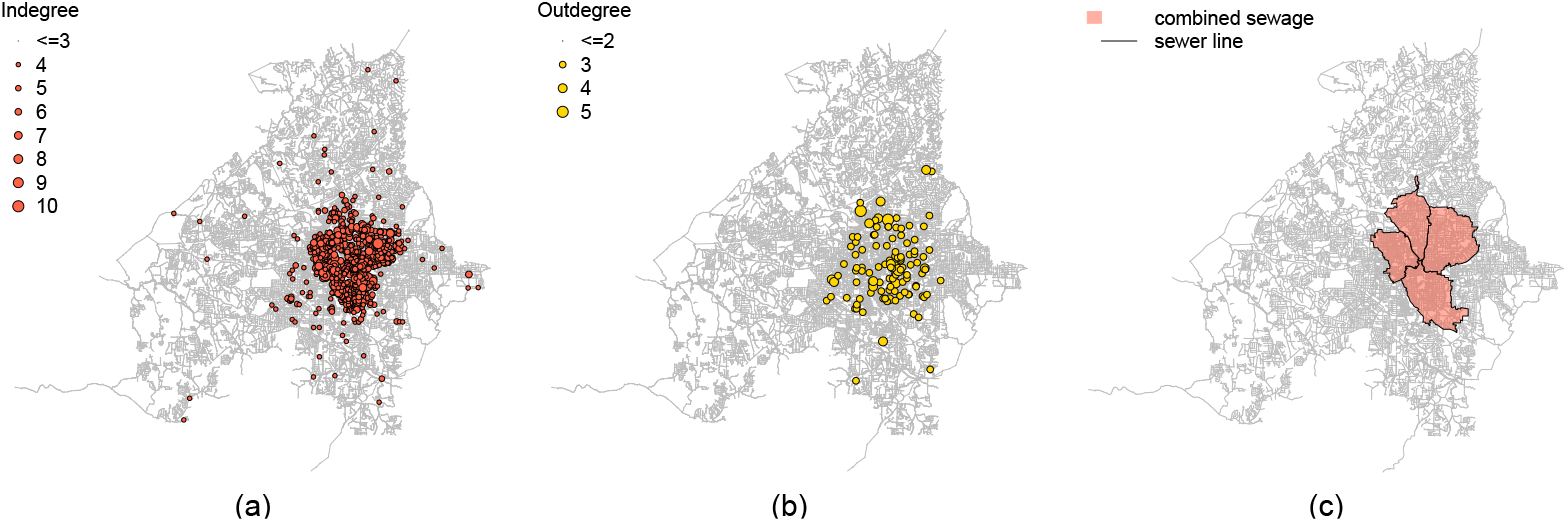
(a) Geographical distribution of manholes with large indegree on the sewer network; Geographical distribution of manholes with large outdegree on the sewer network; (c) Areas with combined sewers in the city of Atlanta.

### Influent Line Wastewater Surveillance Results

A total of 362 samples were collected from nine influent lines entering three wastewater treatment facilities during March 20, 2021–April 11, 2022 (Figure 3a). Figure 4 shows the concentrations of SARS-CoV-2 RNA in the wastewater samples from influent lines increased between July and August 2021, matching the temporal trend of reported cases in Fulton County, Georgia. When the reported cases decreased in October and November 2021, the SARS-CoV-2 RNA concentrations in wastewater stayed relatively high. In late December, the SARS-CoV-2 RNA concentration in wastewater and the reported case numbers surged rapidly and then decreased in January 2022.

**Figure 3:**
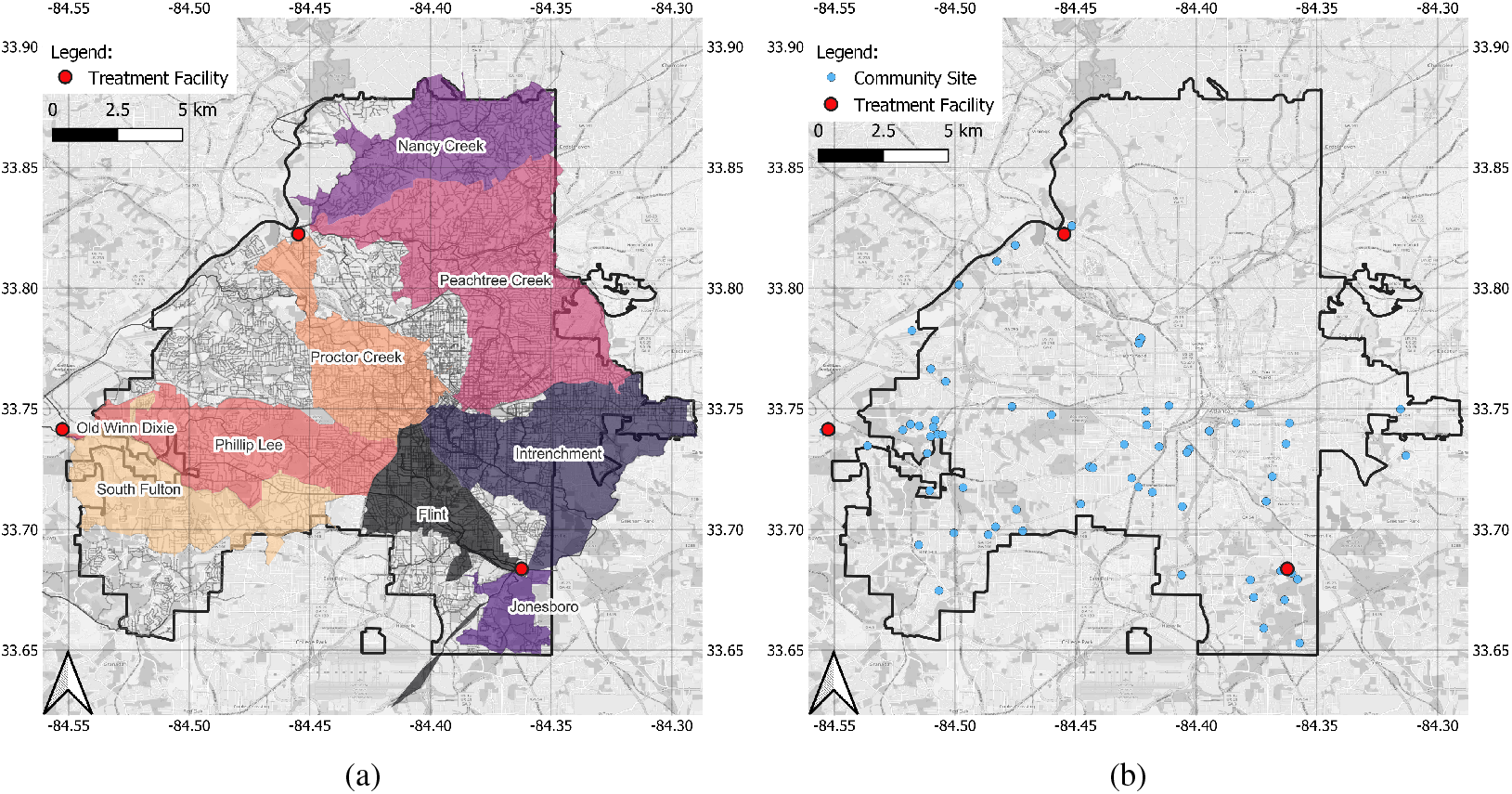
(a) Estimated catchment areas of nine influent lines sites in the city of Atlanta; The catchment area of Old Winn Dixie, which is very small, may not be visible. (b) locations for community sampling sites and school sampling sites.

**Figure 4:**
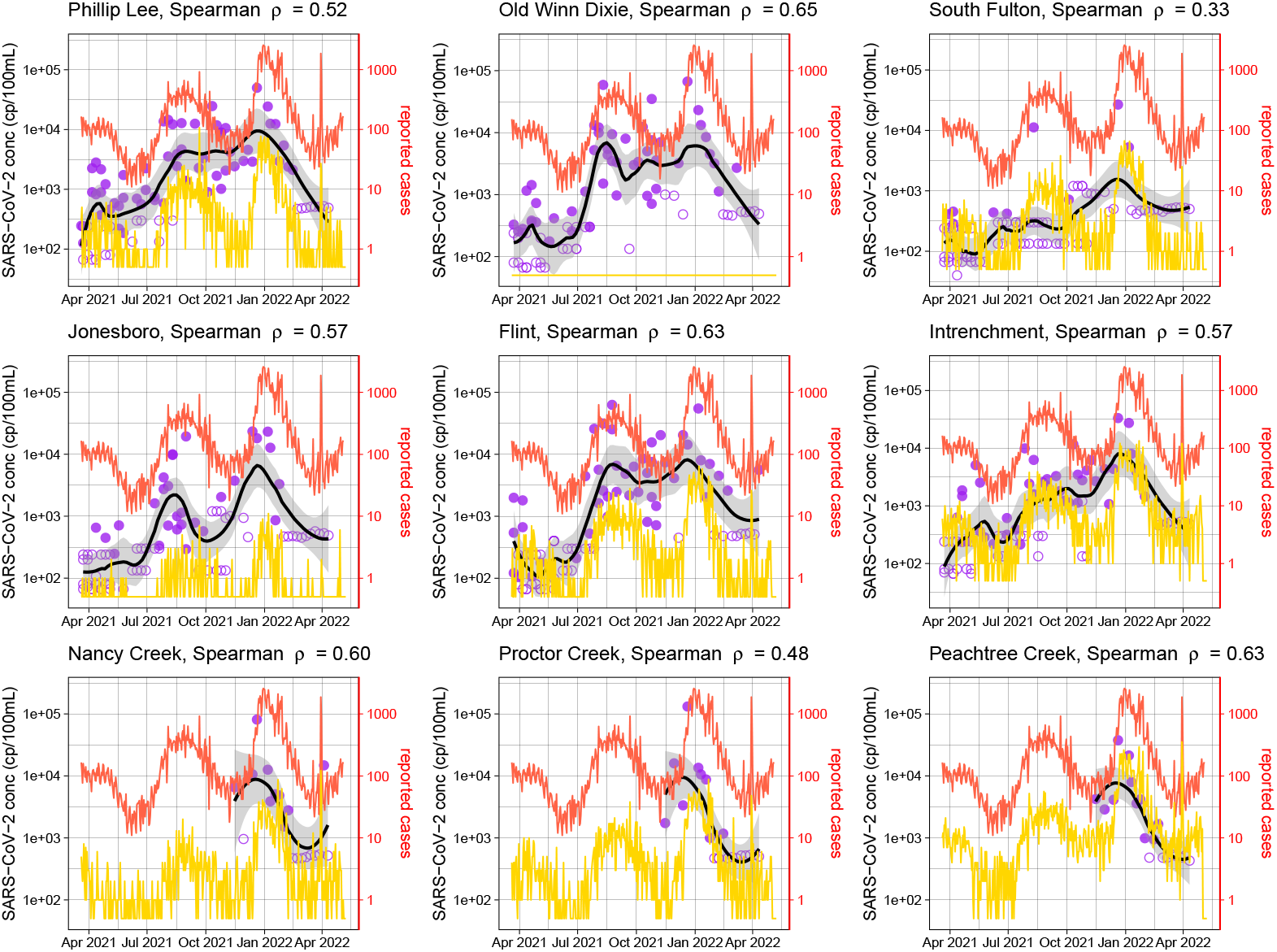
Temporal trends of the concentrations of SARS-CoV-2 RNA in wastewater from nine influent line sites and the reported case numbers during March 20, 2021–May 8, 2022. Solid purple symbols represent positive samples and open purple symbols show negative samples at the detection limit. The black line is the LOESS line for concentration of SARS-CoV-2 RNA, and the gray band represent the 95% confidence interval. The red line is the number of reported cases in Fulton County, Georgia. The yellow line is the number of reported cases in the catchment area of the influent site. Throughout the study period, no case was reported in the very small catchment area of Old Winn Dixie. The graph caption shows the Spearman correlation between the smoothed concentrations of SARS-CoV-2 RNA in wastewater and the number of reported cases in Fulton County.

Correlation analysis showed that the concentration of SARS-CoV-2 RNA in wastewater and the number of daily reported COVID-19 cases in Fulton County were moderately correlated (Spearman ρ between 0.48–0.65) except for the South Fulton site (Spearman ρ=0.33). We also found strong correlations (Pearson’s r between 0.70–0.98 and Spearman ρ between 0.67–0.95) between the SARS-CoV-2 RNA concentrations in the wastewater samples from those six influent line sites sampled for the entire study period (Supplementary Figure 2). For most influent line sites, the correlations between the concentrations of SARS-CoV-2 RNA in wastewater and the reported cases (in Fulton County or in the catchment area) 7–12 days later were higher than the correlation between the concentrations of SARS-CoV-2 RNA in wastewater and the reported cases at the day of wastewater sampling (Supplementary Figures 3 and 4).

### Community Site Wastewater Surveillance Results

Figure 5 shows the overall WWS results from community sampling sites. A total of 506 Moore swab samples were collected from 56 manholes (Figure 3b). In the pilot phase 1 (April–May 2021), we collected wastewater samples from 20 community sites, each for a time period of two weeks. We detected samples positive for SARS-CoV-2 RNA at 7 sites, while the other 13 sites only produced weak positive or negative samples. The detection rates for SARS-CoV-2 RNA at community sites varied even between manholes in close proximity. In phase 2 with the targeted sampling (June–August 2021), the frequency of SARS-CoV-2 detection in the wastewater samples from community sites increased in July–September 2021 along with a surge in number of reported COVID-19 cases in Fulton County. In phase 3 with the nested sampling (September 2021–April 2022), samples from some community sites consistently had detectable SARS-CoV-2 RNA throughout the study period while other sites were only positive in December 2021, when the numbers of reported cases surged to an unprecedented high level. Four community sites (Ruby Harper Blvd, Southside Industrial Parkway, Village Dr, and Walmart), which had low percentages of positive wastewater samples, were relocated at the end of December 2021 to four new sites, which had more positive samples compared to those sites that were dropped. In March–April 2022, when the incidence decreased to a relatively low level, some community sites were still positive for SARS-CoV-2 RNA while their corresponding downstream influent line samples were negative (Figure 6, Supplementary Figures 5, 6, and 7).

**Figure 5:**
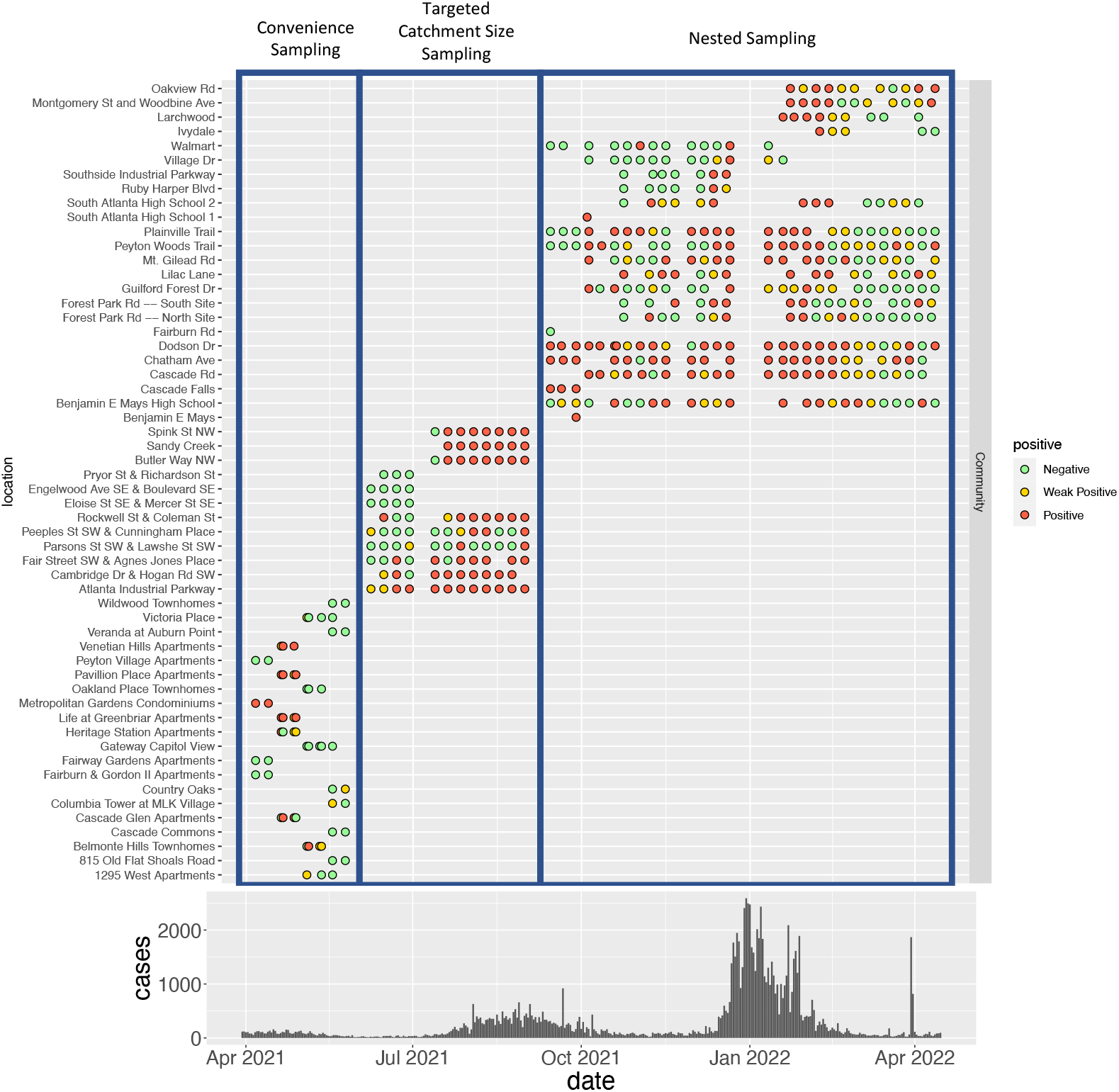
SARS-CoV-2 wastewater surveillance results for community sites and reported case numbers in Fulton County between March 20, 2021–April 15, 2022. The high reported case numbers in April 2022 were caused by a data dump (i.e., delayed reporting), and did not represent a surge in cases during that month.

**Figure 6:**
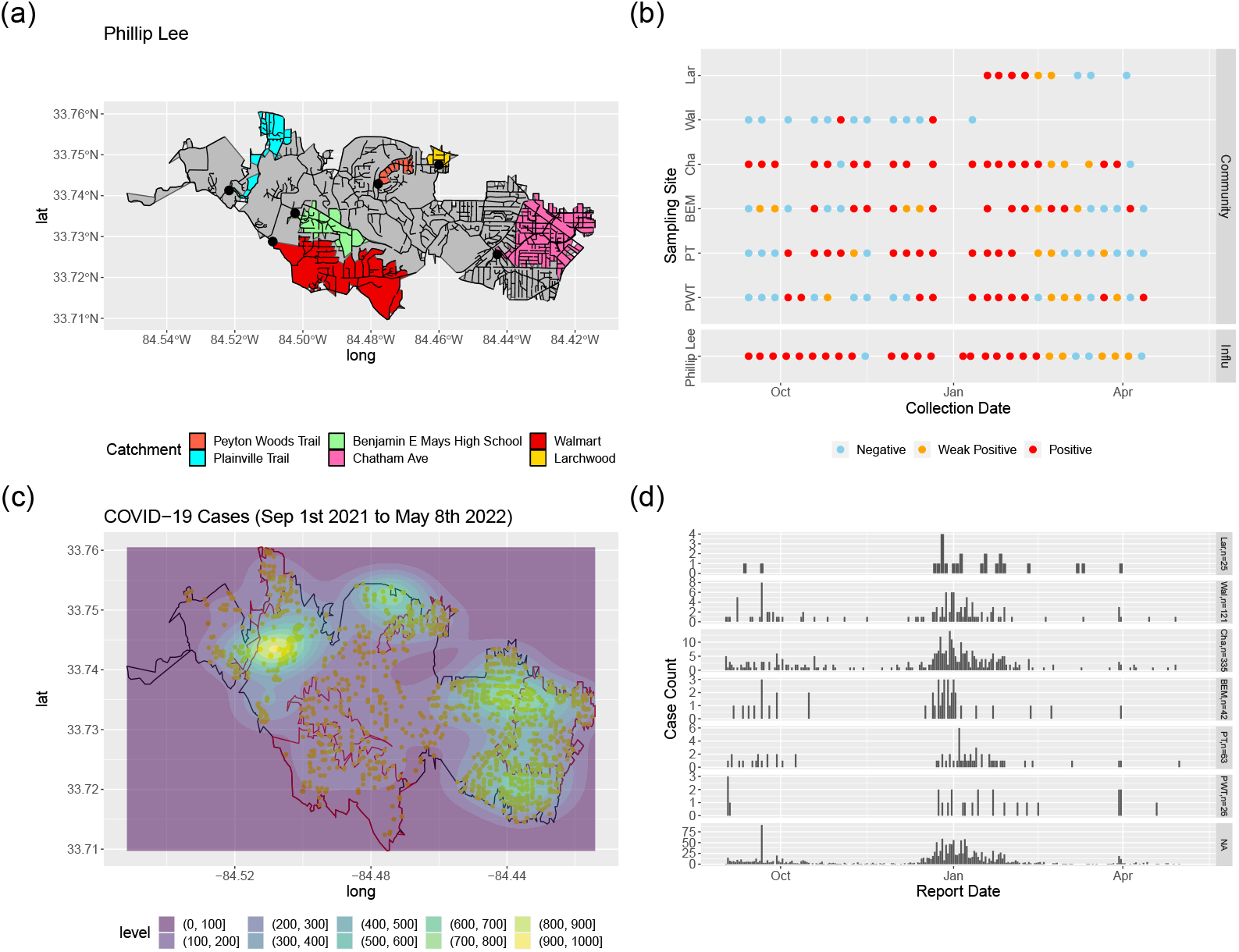
SARS-CoV-2 wastewater surveillance results for community sites and reported COVID-19 case numbers in the catchment area for the Phillip Lee sampling cluster between September 2021–May 2022. Subfigure (a) shows the catchment areas of each community site nested within the overall catchment of the influent line site (in gray). The black lines represent the sewer network lines. Subfigure (b) shows the weekly wastewater surveillance results (RT-PCR detection of SARS-CoV-2 RNA). Subfigure (c) shows the heatmap of reported COVID-19 cases between September 1st 2021–May 8th 2022 within the influent catchment area. Subfigure (d) shows the epidemic curves of COVID-19 within each community site catchment area and NA represents all the cases in the catchment area of the influent line site that are not in the catchment area of a specific community site. PWT, PT, BEM, Cha, Wal, and Lar represent Peyton Woods Trail, Plainville Trail, Benjamin E Mays High School, Chatham Ave, Walmart, and Larchwood respectively.

With the nested sampling design in phase 3, we observed spatial agreement between SARS-CoV-2 RNA detection in wastewater from a community site and the number of cases reported within the catchment area of the same site. Figure 6 shows WWS results and cases reported side by side for the Phillip Lee sampling cluster (one downstream influent line site with multiple upstream community sites) as an example. Catchment areas of community sites with high SARS-CoV-2 detection rates in the wastewater (i.e., Chatham Ave and Plainville Trail) overlapped with areas with high numbers of reported COVID-19 cases. In contrast, samples from the Walmart site, which has a relatively large catchment area, were only positive for SARS-CoV-2 RNA twice out of 13 samples. In this area, few cases were reported during the surveillance period. The results from other sampling clusters are included in the Supplementary Figures 5, 6, and 7.

## Discussion

As new variants emerge, SARS-CoV-2 continues to cause waves of infection across the globe [32]. After more than two years of COVID-19 pandemic, adherence to COVID-19 rules and guidance has declined [33, 34], and there are signs of fatigue in performing PCR tests for individual nasal swabs and reporting daily confirmed COVID-19 case numbers accumulated among patients, caregivers, laboratories, and health departments. Such pandemic fatigue contributes to increasing and more heterogeneous underestimation of COVID-19 incidence all over the world. Meanwhile, it is unrealistic to sustain large scale epidemiological surveillance systems based on individual diagnostic testing given the economic burden especially in countries where the resources are limited. WWS, as an inexpensive, sensitive, and non-intrusive method, could provide information about COVID-19 incidence at different geographic levels and is especially useful for populations where COVID-19 is under-ascertained and/or under-reported due to limited access to diagnostic testing or health behavior. With careful sampling designs, WWS can be utilized to examine temporal trends of COVID-19 incidence at the city level and to identify spatial COVID-19 hotspots at the community level. Such information differentiated by geographic level can directly inform and guide public health responses to COVID-19.

Designing a sensitive and actionable WWS system requires a good understanding of what contributing population or catchment area is represented in a wastewater sample. With a well-characterized sewerage system, the catchment area (and population) can be determined for any sampling site within the sewer network. SARS-CoV-2 RNA concentrations in wastewater collected from such a sampling site reflect ongoing infections in the population who reside in its catchment area. However, sewerage systems usually develop along with the urbanization process. Combined sewers are common in some historical areas, and sanitary sewers are usually added as a city develops and expands. In the city of Atlanta, combined sewers are mainly located in the city center, which is also the most densely populated area of the sewer network. For WWS, sampling at sites from combined sewers poses challenges to interpretation of the results. First, because the combined sewer collects both sewage and stormwater, the concentration of SARS-CoV-2 RNA in the wastewater can be impacted by extreme variation in flow rate and velocity (caused by rainfall) and any chemicals in the stormwater, which may reduce the sensitivity or inhibit the lab assay. In the city of Atlanta, the Intrenchment, Proctor Creek, and Peachtree Creek influent line sites catch large proportions of combined sewers. Second, areas with combined sewers were observed to have many cross-connections (higher connectivity), which may have been designed to avoid blockage and overflow. Catchment areas and populations can not be precisely determined for sampling sites in such sewer networks.

It is critical that catchments of different sampling sites are discrete areas so that the sources of human excreta entering the catchment can be considered independent. Usually, influent lines are separate and distinct before merging at a wastewater treatment facility. Each line typically represents a large proportion of the city. For community manhole sites, any upstream and downstream connection creates challenges interpreting results regardless of whether they are quantified RNA estimates or presence/absence. For example, when upstream and downstream sites are both positive for SARS-CoV-2, we can not determine whether there are COVID-19 infections located in areas covered by the downstream site but not by the upstream site. Independent catchment areas enable a more straightforward approach for linking WWS results with SARS-CoV-2 infections in specific catchment areas (and populations). The selection of independent locations within the network can be achieved with network partitioning methods [29, 30], and in this study we developed a “*main trunk* and *branch sites*” method to achieve this goal. Some limitations are introduced by forcing independence between sites. First, it is challenging to identify independent sites in a sewer network with many cross-connections. The sites selected could have extremely large or extremely small catchment areas. Second, in a tree topology sewer network, a small number of branch sampling sites may only be able to cover a proportion of the sewer network. A careful selection of *branch sites* (multiple sites as a system) should be located on a tree topology sanitary sewer network, have independent catchment areas, and together capture wastewater inputs from as large an area of interest as possible.

At the city level, routine COVID-19 WWS, with weekly or multiple samples per week, could be conducted as a supplemental or alternative method of epidemiological surveillance to provide incidence information for large areas of a city. Although estimating the number of COVID-19 cases or SARS-CoV-2 infections (symptomatic and asymptomatic) directly from wastewater results involves many factors [35], the time course of SARS-CoV-2 RNA level in wastewater should reflect the time course of numbers of viral shedders in the population connected to the sewerage system. Similar to previous studies [36, 37, 38, 39], this study found temporal correlations between SARS-CoV-2 RNA concentrations in wastewater and numbers of daily COVID-19 cases reported. With weekly sample collection, we observed a 7–12 days lead time in wastewater signal trends in influent line samples compared to trends in reported cases in the catchment areas for these influent line sites. This finding demonstrates the potential of WWS to provide early warning of COVID-19 case surges in the city, which could forecast needs for clinical and diagnostic testing resources. However, there was also some discordance between the trends of the SARS-CoV-2 RNA concentration in wastewater and the number of reported COVID-19 cases. Many wastewater samples from the South Fulton influent line were negative for SARS-CoV-2 RNA even when there was a large number of reported cases in the catchment area. We have been exploring the reasons for this finding by measuring pH values, suspended solids, and turbidity of all the wastewater samples from influent lines since July 2021. Higher pH values (>10) were frequently measured (68%) in the wastewater samples collected from the South Fulton influent line compared to pH values (around 7) in the wastewater samples from other influent lines. High pH may denature the virus and degrade the viral RNA, and it may also indicate the presence of other chemicals in wastewater that may cause PCR inhibition. We also observed sustained high SARS-CoV-2 RNA concentrations in some influent line samples after the reported COVID-19 case numbers declined, which could be caused by prolonged fecal shedding [40] or may indicate continuing silent transmission in some communities. The SARS-CoV-2 RNA concentration in wastewater may be considered to represent the number of infected people currently shedding the virus in their feces, rather than COVID-19 incidence. Therefore, concentrations of SARS-CoV-2 RNA in wastewater can be considered a distorted reflection of the reported numbers of COVID-19 cases, convoluted by the fecal shedding curve which causes delay and averages over a certain time period. This distortion weakens the correlation between concentrations of SARS-CoV-2 RNA in wastewater and numbers of COVID-19 cases reported. Silent transmission occurs when an asymptomatic case passes the virus to someone else. As the pandemic continues, there will be more people with some level of immunity, who may be infected with mild or no symptoms. Those infections, which may be not captured by epidemiological surveillance based on diagnostic testing, are still contributing to the disease transmission and are shedding SARS-CoV-2 in their feces. In such a scenario, WWS is valuable to guide public health response especially in settings where resources are limited or there is severe underreporting/under-ascertainment.

Sampling wastewater from upstream community sites in addition to sampling from downstream influent sites (i.e., nested sampling) improves the sensitivity of wastewater-based surveillance system. SARS-CoV-2 RNA signals at downstream sites could be subject to large dilution, rapid degradation of virus and viral RNA, and substantial loss of signal while moving through the sewer lines [15]. In this study, while the samples from South Fulton influent line site frequently failed to detect SARS-CoV-2 potentially due to high pH values, wastewater samples from its upstream community sites consistently tested positive for SARS-CoV-2 RNA. In this type of situation, sampling only from the downstream influent line site produced false negative results. And when the disease incidence is low, the concentration of SARS-CoV-2 RNA could be diluted to a level below the limit of detection whereas sampling at the upstream sites, that are closer to the shedders, could be more sensitive [15].

With an estimated over 413 million people infected with SARS-CoV-2 and 4.27 billion people fully vaccinated worldwide [41], immunity has gradually increased in the population. However, acceptance and availability of COVID-19 vaccines are not uniformly distributed [42] resulting in more heterogeneous and localized COVID-19 transmission [43]. Given these circumstances, more targeted prevention and control measures are needed to allocate the resources (testing capacity, vaccines, health communications, and healthcare resources) to those communities at risk. Reported COVID-19 cases can be geocoded with the assistance of other existing data systems (e.g., voting registration, vehicle registration, medical records etc.) to identify spatial clustering of cases. However, geocoding all reported cases is not a standard procedure in the epidemiological surveillance and requires tremendous effort. In this study, we evaluated the feasibility of using WWS at the community level to trace disease hotspots in a spatial area. With a nested sampling design, *branch sites* that cover high COVID-19 incidence areas tend to have high SARS-CoV-2 RNA detection rates. This study demonstrates how such spatial coincidence enables WWS to be deployed to locate infection clusters.

Community level WWS with a small number of independent sampling sites usually cannot cover the entire geographic area. The adaptive sampling method in the current study allows relocation of sites not detecting SARS-CoV-2 and searching for hotspots in the area. Furthermore, with immunity waxing and waning in the population, and the periodic introduction of new SARS-CoV-2 variants, disease transmission will continue to vary both temporally and spatially. For example, populations in COVID-19 hotspots during the latest Omicron wave (December 2021–February 2022) may gain some level of immunity and be less likely to contribute to SARS-CoV-2 transmission for several months afterwards. Then, the COVID-19 hotspots for the next wave of infection could move to areas where the population has a low level of immunity. An adaptive sampling approach can transform WWS into a dynamic system to identify and trace COVID-19 hotspots in communities.

This study highlights the importance of carefully designed sampling strategies for developing a sensitive and sustainable WWS. We have developed, applied, and validated a novel nested sampling strategy with adaptive sampling process, which enables identification and tracing of COVID-19 hotspots. The strategic sampling design described here is critical for long-term sustainability of WWS for COVID-19 and other diseases, and provides maximum value of information from a minimum number of samples. The spatial granularity of this wastewater surveillance data provides actionable information to guide COVID-19 prevention and control measures at different geographic levels.

## Methods

### Study Area

The current study was conducted in the city of Atlanta, which is the capital and most populous city (498,715 in 2020) within the state of Georgia. The City of Atlanta Department of Watershed Management (DWM) manages the wastewater system in Atlanta and serves 1.2 million customers within the Atlanta Metropolitan Area. There are three water reclamation centers (WRCs) within the city limits of Atlanta, and each WRC has multiple influent lines (Figure 3a). South River WRC has a permitted treatment capacity of 48 million gallons per day (MGD) and provides wastewater treatment for portions of Atlanta, East Point, Hapeville, College Park, and parts of DeKalb County and Clayton County. Utoy Creek WRC has a permitted treatment capacity of 40 MGD and provides wastewater treatment for portions of Southwest and Northwest Atlanta, East Point, and Fulton County. R.M. Clayton is the largest among the three WRCs, with a permitted treatment capacity of 100 MGD. R.M. Clayton provides wastewater treatment services for the City of Atlanta, primarily north of Interstate 20, a portion of Sandy Springs, and Northern DeKalb County. Detailed information on the WRCs and the sampled influent lines can be found in Supplementary Table 1. The community sampling sites in this study were selected mainly in the southern part of the city (south of Interstate 20), where underserved neighborhoods (e.g., Adamsville, Oakland City, Pittsburgh, Mechanicsville, and the West End) are located (Figure 3b). The Georgia Department of Community Affairs defined underserved areas as areas with the highest unemployment rate, the lowest per-capita income and the highest percentage of residents whose incomes are below the poverty level. People living in the underserved areas, when infected by SARS-CoV-2, are less likely to seek health care and get tested, leading to higher levels of COVID-19 under-reporting and under-ascertainment via traditional epidemiological surveillance.

### Sewer Network and Catchment Identification

Once the SARS-CoV-2 viruses in feces enter sewage, they are transported through the sewerage system consisting of sewers (gravity sewers and force mains), manholes, pumping stations, and wastewater treatment facilities. A sewerage system can be viewed as a flow network with manholes (potential sampling sites) as nodes, sewer segments as edges, and wastewater treatment facilities as sinks which only have incoming flows. The directions of edges represent the flow direction of sewage in sewer lines. If pumping stations are not involved, the sewage flow is only driven by gravity, and the flow direction is determined by the elevation.

A simplified flow network (e.g., rivers, sewerage system) could have a line, tree, or net topology. In a line topology network, the water flows from one upstream point (node) to one downstream point without merging or splitting (Figure 7a). The indegree (the number of edges entering the node) and outdegree (the number edges leaving the node) both equal 1 for all the nodes in the network. A tree topology network has multiple upstream branches (edges) that merge as they flow downstream (Figure 7b). The indegree could be larger than 1 for some nodes while the outdegree equals 1. The larger the average indegree of nodes, the more branches exist within the tree network. In a net topology network, the flow could split and merge multiple times as the water travels downstream (Figure 7c). The indegree and outdegree both are larger than 1. The larger the indegree and outdegree, the higher the connectivity within the network. A sewer network has a hybrid topology integrating line, tree, and net topologies. Usually, sewage is collected from geographically widespread points and moves to the final destination of a wastewater treatment facility. In this type of network, the majority of nodes have outdegrees equal to 1, and the sewer lines (edges) are merging, developing mainly a tree topology. In this study, we examined the network statistics (indegree and outdegree) and network topology of the Atlanta sewer network using geospatial data for the Atlanta sewerage system (with flow direction), in shapefile format, provided by the DWM.

**Figure 7:**
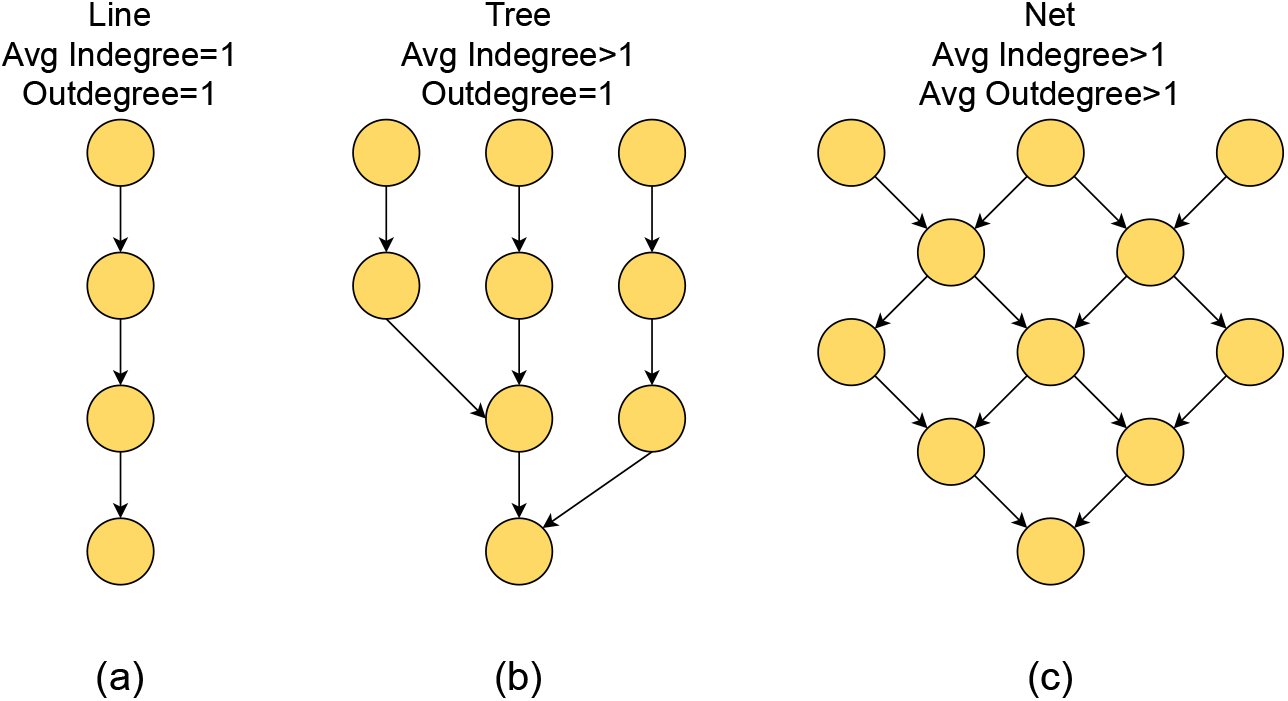
Indegree and outdegree of different flow network topologies.

For WWS, it is critical to understand the geographic area and approximately how many people are represented in each wastewater sample. Based on the sewer network data, we identified all the upstream points (nodes) and sewer lines (edges) for any potential sampling site (manhole) in the sewer network using “lucy” [44], a wrapper for the igraph package in R. In the current study, such an upstream sub-network was defined as the catchment of a sampling point. The catchment area was estimated as the polygon area covering this sub-network using the “concaveman” package [45] in R, and the catchment size was approximated by information on the number of upstream manholes (including the sampled manhole).

### Sampling Design and Sample Collection

One of the main purposes of this WWS study was to monitor COVID-19 incidence in the city of Atlanta at a city level. In this study, the incidence was defined as the number of new COVID-19 cases reported to the Georgia Department of Public Health (GDPH) over a specific period of time. During March 2021–April 2022, weekly 1 L grab samples of wastewater were collected on Monday mornings at six influent lines of two water reclamation centers (Utoy Creek and South River), identified by partners at the DWM. Since November 2022, samples were also collected from three additional influent lines at the R.M. Clayton WRC. The samples from each influent line represent large areas of the city (Figure 3a) and were intended to monitor temporal trends of incidence within each catchment area. To further assess COVID-19 incidence in smaller geographic areas, samples were collected from upstream community sites at manholes. Unlike wastewater from the influent lines, upstream wastewater is less likely to contain SARS-CoV-2 RNA. The Moore swabs method [46], a low-cost composite sampling approach, was used for community site sampling. During April 2021–April 2022, Moore swabs were placed weekly in manholes across the city of Atlanta (Figure 3b) and retrieved after approximately 24 hours. The details of site identification, sampling requirements, sampling schedule, and the mobile data collection process are described in Supplementary Material A.

We developed the Atlanta WWS sampling design for community sites in three phases. In the pilot phase 1 (April–May 2021), information related to the sewer network was not available. We conducted convenience sampling by selecting community sampling sites (manholes) in low-income neighborhoods in South Atlanta based on insights from our partners at DWM. The sampling sites were relocated to new sites every two weeks to explore sampling in different areas of the city and examine the variation in detection rates at different community sites. In phase 2 (June–August 2021), we obtained the Atlanta sewer network shapefile from the DWM, which enabled us to identify the catchment areas and catchment sizes for all manholes. Based on the catchment sizes (measured as number of manholes upstream of the target manhole), we were able to select sampling sites with small, medium, and large catchment sizes (Supplementary Table 2). Three community sites with low detection rates were relocated after four weeks. In phase 3 (September 2021–April 2022), we set up a nested sampling design by sampling one downstream influent line site along with multiple upstream *branch sites* (community manholes), which was defined as a sampling cluster. Using the Phillip Lee sampling cluster as an example (Supplementary Figure 1), we identified the *main trunk*, which was the longest path in the sewer network with the influent line site as the end point. Tributary *branch sites* were defined as the points (nodes) before merging into the *main trunk*. The *branch sites* were independent of each other (no upstream and downstream relationship), and the initial goal was to cover as many manholes as possible. Independence between different *branch sites* is critical to identify sub-areas with high incidence. In phase 3, we also applied the adaptive sampling process [15] to relocate community sites within the nested sampling design in January 2022. Sites that were rarely positive for SARS-CoV-2 RNA were replaced with new *branch sites*, which were proposed following the descending order of their catchment sizes (largest to smallest).

### Adaptive Sampling Process

Usually, the independent upstream sampling sites with small catchment areas could not cover the entire influent line site catchment area that we wanted to monitor. Some high incidence areas, if not covered, could be undetected by WWS. The adaptive sampling process relocated a small proportion of sites periodically based on the most recent WWS results collected. The adaptive sampling process has the following steps:

#### Initialization

Assuming we are designing nested sampling on a sewer network and *n* sampling sites are planned, one sample site will be located at a downstream location of the sewer network (end point of sewer network), and *n* −1 branching sites (before entering the *main trunk*) will be selected based on their cumulative weight. Each of the manholes (nodes) was given a weight as 1 at initialization. The cumulative weight for each manhole is calculated by adding the weights of all the manholes upstream of the target manhole. The *branch sites* are selected as independent (no upstream and downstream relationship between *branch sites*) sites with the largest cumulative weights.

#### Weighting

Once the sampling sites have been initialized, wastewater is sampled and tested for a couple of rounds. When the presence of SARS-CoV-2 RNA (positive) is detected in samples from a specific site, the weights of manholes within the catchment of this site increase. The percent that the weight increases depends on the proportion of positive samples. On the other hand, if no positive result is detected in samples from a specific site, the weights of manholes within the catchment of this site decrease.

#### Updating

After the weights of manholes have been adjusted, the *branch sites* are re-selected following the process same as initialization but based on updated cumulative weights. The weighting and updating steps are repeated periodically, and the WWS can be transformed into a dynamic system that updates itself based on the most recent results.

Figure 8 illustrates the application of the adaptive sampling process in a simulation study. The hypothetical high-risk area is located at the left top corner of the sewer network. We initialized the sampling with one site at the end of the sewer network and five branching sites. The weighting and updating steps were conducted after every two rounds of weekly data collection. As the update process went on, one or two sampling sites were relocated and the weight of the manholes (represented as the size of nodes) on the left top corner increased. The source code of the adaptive sampling process can be found on Github (https://github.com/YWAN446/COVID-WWS-ATL).

**Figure 8:**
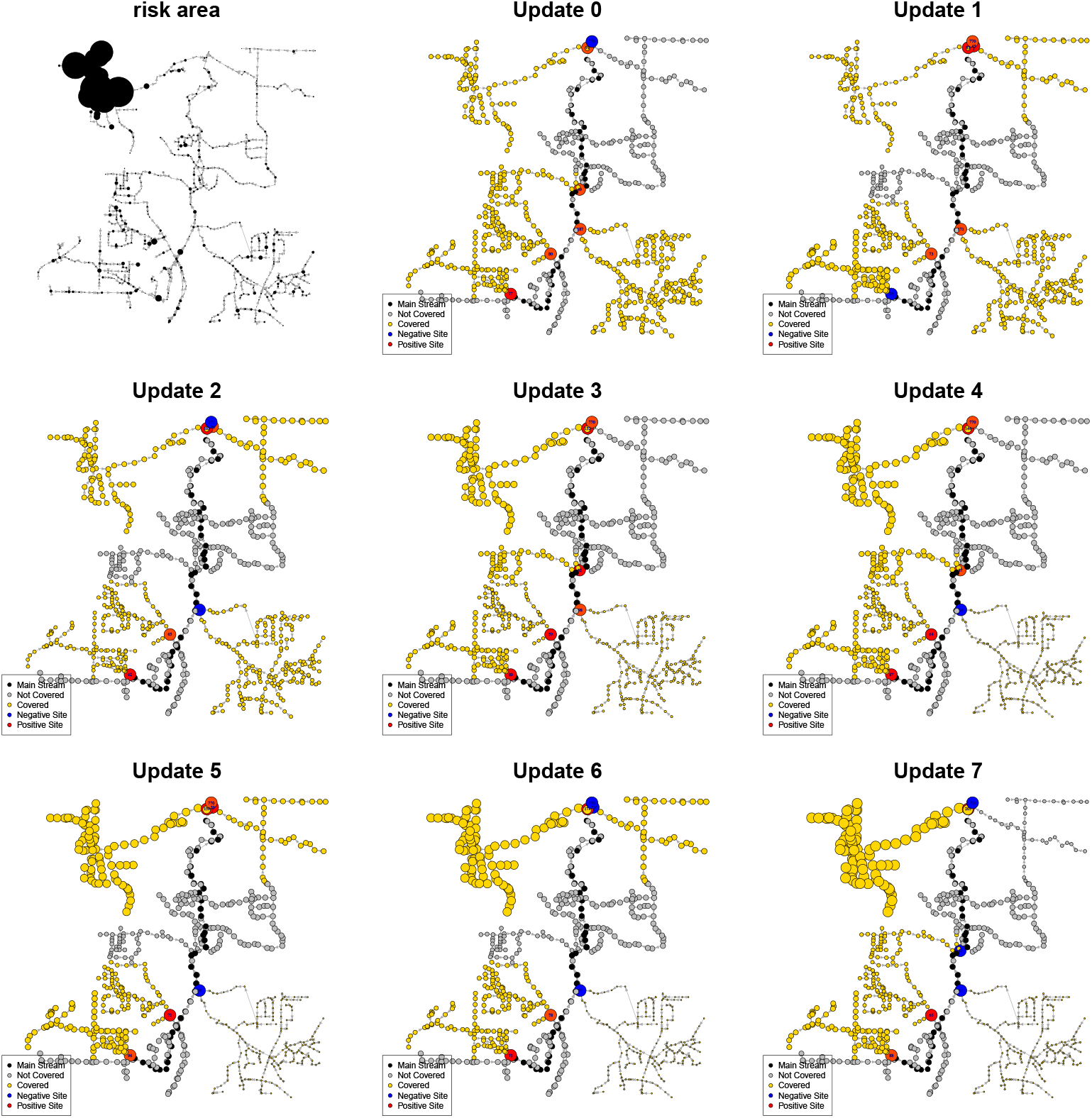
Illustration of adaptive sampling process in a simulation study. Each node is a manhole in the sewer network. Manholes shown in black are on the longest line in the sewer network, defined as the *main trunk* (main stream). Manholes shown in yellow are covered by the selected sampling sites, while manholes shown in gray are not covered. Red manholes represent sites with positive wastewater samples (detection of SARS-CoV-2). Blue manholes represent sites where the wastewater samples were negative. The sizes of nodes represent the weights of sites. The larger the weight, the higher estimated risk of COVID-19 transmission.

### Sample Processing and Lab Testing

Between March–October 2021, three methods were used to concentrate SARS-CoV-2 from wastewater samples and extract viral RNA: (1) The Membrane Filtration method, which incorporates the use of a 0.45 µm membrane filter, was used to capture SARS-CoV-2 from wastewater grab samples. The Qiagen RNeasy Mini kit (Qiagen, Germany) was used for RNA extraction [46]. (2) The Skim Milk method was used to concentrate of SARS-CoV-2 in Moore Swab samples and the same Qiagen RNeasy Mini kit was used as the Membrane Filtration method (dx.doi.org/10.17504/protocols.io.b2uwqexe). (3) The Manual Nanotrap® Concentration method involved CERES Nanotrap® magnetic hydrogel particles and an enhancement reagent (Ceres Nanosciences Inc., USA). The Nanotrap® Magnetic Virus Particles capture and bind the SARS-CoV-2 viral particles in the wastewater so that they can be concentrated into a smaller volume. The Qiagen QIAamp Viral RNA Mini kit was used for RNA extraction (dx.doi.org/10.17504/protocols.io.b2uzqex6). The transition between these three methods to our final automated KingFisher Apex system occurred in October 2021 and the MagMax Viral/Pathogen Nucleic Acid Isolation kit (Thermo Fisher Scientific, USA) was incorporated in the KingFisher platform (dx.doi.org/10.17504/protocols.io.b2nkqdcw). Each protocol mentioned above utilized a sample processing control of Bovine Respiratory Syncytial Virus (BRSV) (MWI Animal Health, USA), which was spiked directly into wastewater samples prior to sample processing. After concentration and RNA extraction, SARS-CoV-2 and BRSV were detected by a singleplex real-time quantitative reverse transcription polymerase chain reaction (RT-qPCR) (dx.doi.org/10.17504/protocols.io.b2qyqdxw) between March–November 2021. After December 2021, a duplex RT-qPCR platform for simultaneous detection of SAS-CoV-2 and BRSV was used between December 2021–April 2022. The details for the RT-qPCR methods were described by Liu et al. [46]. The result was classified as positive when both CT values were present from the duplicate wells and at least one CT value was below 36. If both CT values were present and above 36, the sample was considered as weak positive. When any CT from duplicate wells was absent, the sample was considered negative. For grab samples, the concentrations of SARS-CoV-2 RNA in wastewater were estimated from CT values using standard curves.

### Comparing Wastewater Surveillance Results with Reported Case Data

COVID-19 cases reported to the GDPH from March 2021 through April 2022 in Georgia were geocoded using Esri’s ArcGIS Streetmap Premium location information for the North America region by GDPH. A set of data sources were used with resident address data from the case report form having the highest priority, followed by electronic laboratory records, and administrative data from other agencies serving Georgia’s residents. Coordinates from geocoded resident addresses were used if they passed a series of quality score measures (e.g., resident address matched a single address with the highest score). Records not meeting these quality criteria were excluded. The final data file was the most complete and precise representation of resident address for COVID-19 cases in Georgia.

Wastewater results from any sampling site were matched with a subset of geocoded COVID-19 cases located within the catchment area of the sampling site. For each influent line site, we examined the temporal trends of SARS-CoV-2 RNA concentrations in wastewater and case numbers reported in the catchment area. The concentrations of SARS-CoV-2 RNA were smoothed over time using the LOESS (locally estimated scatterplot smoothing) method. The smoothed concentrations of SARS-CoV-2 RNA and the numbers of daily reported COVID-19 cases in Fulton County/catchment area were analyzed for correlation and temporally lagged correlation. For community sites, we examined the spatial agreement between catchment areas of wastewater sampling sites with high SARS-CoV-2 detection rates and high COVID-19 incidence areas from the geocoded case data. The wastewater detection rate of a site was calculated as the percentage of positive samples and heatmaps of COVID-19 cases were generated using 2D kernel density estimation. Data analysis and data visualization in this study were conducted using R 4.0.1. The maps in Figure 3 were generated by ArcGIS.

The GDPH Institutional Review Board determined that this analysis was exempt from the requirement for IRB review and approval and informed consent was not required.

## Data Availability

The dataset of WWS results is available at https://github.com/YWAN446/COVID-WWS-ATL/data. The geocoded COVID-19 case data used in this study is available through the Public Health Information Portal data request process (https://dph.georgia.gov/phip-data-request).

## Code Availability

The codes for performing the data analyses, data visualization, adaptive sampling process, and simulation study are available at https://github.com/YWAN446/COVID-WWS-ATL.

## Acknowledgements

This project was funded by the NIH Rapid Acceleration of Diagnostics (RADx) initiative with federal funds from the National Institute of Biomedical Imaging and Bioengineering, National Institutes of Health, and the Public Health and Social Services Emergency Fund through the Biomedical Advanced Research and Development Authority, HHS Office of the Assistant Secretary for Preparedness and Response, Department of Health and Human Services, under Contract No. 75N92021C00012 to Ceres Nanosciences. We thank all the support from Salvins J. Strods and Chipo Chemoyo J. Baker Afame-funa from RADx Initiative, and Dr. Louis J. Vuga from NIH. The data collection was conducted and supported by the City of Atlanta Department of Watershed Management team, including Deputy Commissioner Quinton Fletcher, Brantley Doctor, Elizabeth Sanchez, Batsirai Nyandebvu, Shatoya Hinkle. We appreciate the discussions with partners at the Georgia Department of Public Health, including Dr. Laura Edison, Dr. Amanda Feldpausch, Cristina Meza, John Olmstead, and Pravara Harati. We appreciate all of the contributions made to this project by colleagues at Ceres Nanosciences, including Ross Dunlap, Ben Lepene, Dr. Robbie Barbero, and Tara Jones-Roe, who dedicated tremendous effort during the early days of the COVID-19 pandemic to manufacture the Nanotrap Magnetic Virus Particles and rapidly develop an easy-to-use and powerful set of methods for SARS-CoV-2 RNA concentration from wastewater. This study was also supported by Dr. Allison Chamberlain and Eve Rose from Emory COVID-19 Response Collaborative (ECRC). We thank all the Emory students involved in the study: Stephen Mugel, Matthew Cavallo, Caleb Cantrell, Jillian Dunbar, Makoto Ibaraki, Kelly Geith, Lindsay Saber, Rebecca Kann, Keyanna Ralph, Haisu Zhang, Lutfe-E-Noor Rahman, and Weiding Fang.

## Author Information

### Contributions

Y.W., P.L., M.W., M.B., P.FM.T., and C.L.M. conceived study. P.L., J.V., L.G., O.S., L.F., W.R., C.H., and M.B. collected data. Y.W., S.P.H., and P.FM.T. developed the adaptive sampling model and analyzed data. All authors interpreted results and wrote paper.

## Supplementary Materials

## A. Sampling Design and Sample Collection

### A.1. Site Identification

#### Wastewater Treatment Facilities (WWTFs)

The City of Atlanta Department of Watershed Management (DWM) routinely collects samples from influent lines of WWTFs for industrial pre-treatment wastewater monitoring. With the consultation of operators at WWTFs, partners at the DWM identified sampling access points for three main influent lines entering each WWTF.

#### Community Sites

After manholes of community sites were identified using information on the sewer network, the manholes were screened for accessibility in the field. The manholes needed to be accessible and not located in an area with busy traffic (e.g., busy roads) or far from the road (e.g., in the woods). For manholes in grassy or wooded areas, risks were considered for trip hazards (e.g., rocks, sticks, and uneven footing), wildlife (e.g., ticks and snakes), and strenuous hikes. When a manhole was not accessible, the enumerator traced upstream to find an alternative manhole that was accessible.

### A.2. Sampling Requirements

#### Permission

Wastewater sample collection was conducted cooperatively by the Emory University sampling team and the DWM team. The DWM has permission to access and sample from manholes within their jurisdiction (i.e., within the city of Atlanta). For the Emory team to help with the sample collection process, they were required to obtain City of Atlanta contractor badges which were worn in the field. The contractor badges also granted access to the WWTFs without the need for an escort from the DWM team. ***Equipment***: Depending on the type of samples (grab vs. Moore swab) that were collected, different types of equipment were brought into the field. For collecting grab samples, 1 L autoclavable bottles, metal bucket (with handle) and rope, and long-handled water sample dipper or a painter’s pole rigged with a “seat” for the 1 L bottle to sit in were used. For collecting a Moore swab sample, fishing lines (weighted for 50-lb), cotton gauze, bendable metal (e.g., metal coat hanger), a magnetic hook, thin ropes, and a collection bag (e.g., quart-size Ziploc bag, WhirlPak, Biohazard Specimen Transport Bag, etc.) were used. In addition, tape (for labeling collection bottles), a permanent marker, a cooler, ice or ice pack(s), and personal protective equipment (e.g., disposable gowns and gloves, N-95 mask, and face shield etc.) were used for sample collection as needed. A protocol that describes the materials and methods used in the field to collect Moore swabs and grab samples for wastewater sample collection was published on protocols.io (http://dx.doi.org/10.17504/protocols.io.b2rzqd76).

#### Travel Time

For Atlanta COVID-19 wastewater surveillance, travel time between sites typically ranged from 10 to 30 minutes, cumulatively 1–3 hours of driving per day, which includes the travel time between the laboratory at Emory campus and the sampling sites.

#### A.3. Sampling Schedule

#### WWTFs

The DWM team collected influent line samples from Utoy Creek Water Reclamation Center and South River Water Reclamation Center. These two plants primarily serve low-income populations in Fulton County. The Emory sampling team collected samples at influent lines of R.M. Clayton plant, which primary serves high-income populations in Fulton County. Grab samples were collected from these nine influent lines every Monday morning, stored in a cooler on ice, and delivered to the laboratory on Emory campus by noon.

#### Community Sites

The DWM team collected Moore swab samples from manholes located within multiple low-income neighborhoods in South Atlanta. Moore swabs were placed in the wastewater stream of ten manholes on Monday mornings, retrieved on Tuesday mornings, and delivered to the laboratory by 1 PM on Tuesday. Beginning in November 2021, the DWM team began collecting Moore swabs at six additional sites over the weekend (placed on Saturday mornings and retrieved on Sunday mornings). After swabs were retrieved on Sunday, they were stored overnight in a refrigerator at 4 ^◦^C, picked up on Monday mornings at 7:30 AM, and delivered to the laboratory by 8:00 AM.

### A.4. Mobile Data Collection

The DWM GIS team utilized Esri’s Field Operations Apps, including ArcGIS Workforce, ArcGIS Field Map, and ArcGIS Survey123, to conduct the field data collection. ArcGIS Workforce allowed sampling teams to be assigned specific manholes to collect samples from thus avoiding confusion if multiple manholes were present in the area. A dispatcher created weekly sampling assignments from ArcGIS Workforce’s web-based map. The sample collection teams used ArcGIS Workforce’s mobile app to view assignments, get driving directions, and open ArcGIS Field Maps mobile app. ArcGIS Field Maps allowed teams to view and interact with City of Atlanta’s sewerage system. A custom URL was created and linked to a form in ArcGIS Survey123. The custom URL was configured to communicate attributes of the sampled manhole (e.g., manhole ID, pipe size etc.) to ArcGIS Survey123 and avoid teams having to copy and paste data. Collection date and time, team members ID, and geographic location information were automatically populated when the form in ArcGIS Survey123 opened. Teams then filled out a few questions, took photos of sampling site, and scanned the barcodes on the bottles or collection bags. The barcode number was then populated in the form. The teams submitted the forms in the field using cellular data and then continued to the next assigned sampling site. Later, the teams entered the date and time the bottles were delivered to the lab at Emory University for testing. ArcGIS Survey123 created a point feature in a feature layer in ArcGIS Online. These feature layers were analyzed in ArcGIS Pro and exported in multiple data formats. Results from the lab were then populated in ArcGIS Online. ArcGIS Dashboards used the feature layer to display the data. Widgets in ArcGIS Dashboards were configured to sort the data by date, council district, and neighborhood planning unit (NPU). The integration between multiple apps and the seamless transfer of data is the reason why Esri Field Operations Apps were chosen. The required training was minimal due to the intuitive work flow of the apps.

**Supplementary Figure 1.**
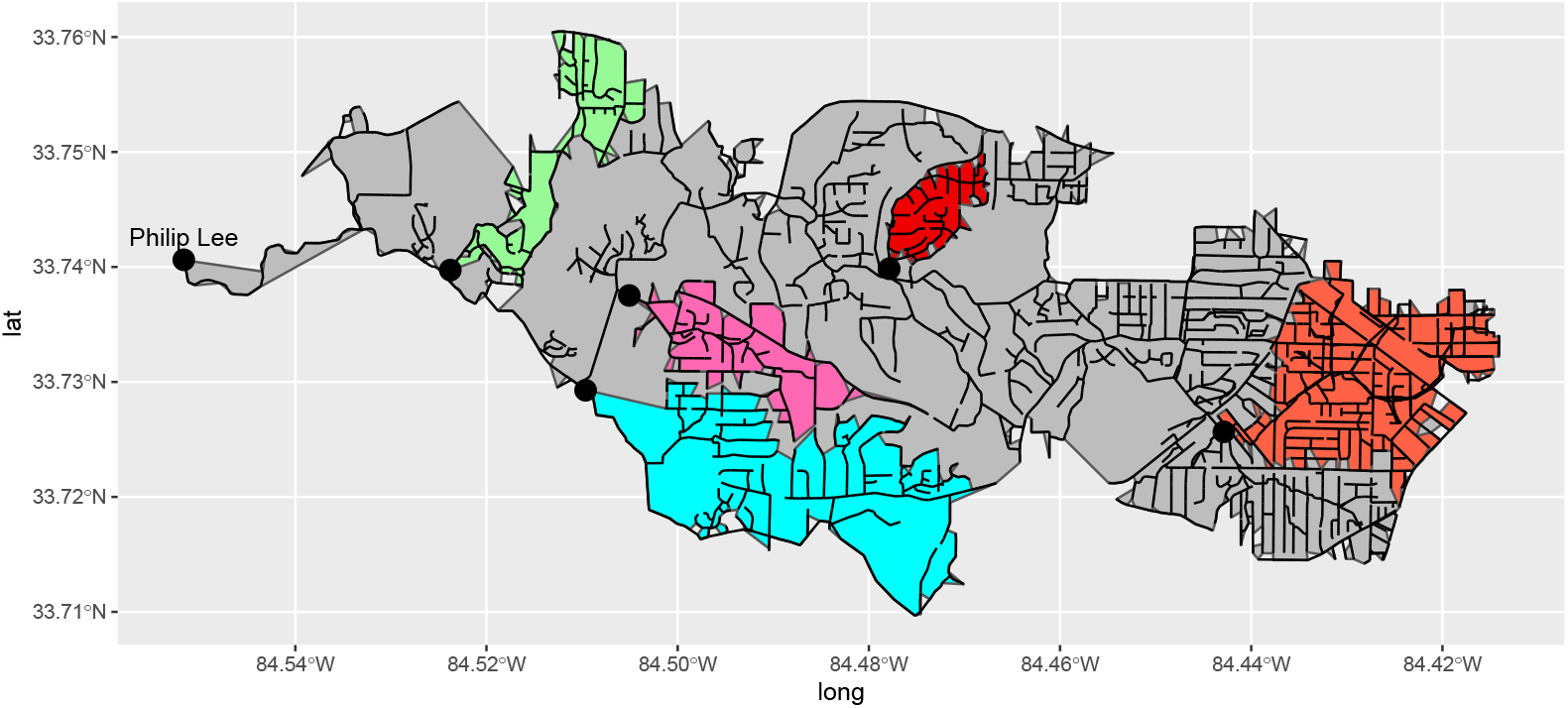
Illustration of nested sampling design for the Phillip Lee sampling cluster, including one influent line site and five independent community sites nested within the catchment area of the Phillip Lee influent line site. The gray polygon represents the catchment area of the Phillip Lee influent line site. The five polygons with different colors represent catchment areas of specific community manhole sites.

**Supplementary Figure 2.**
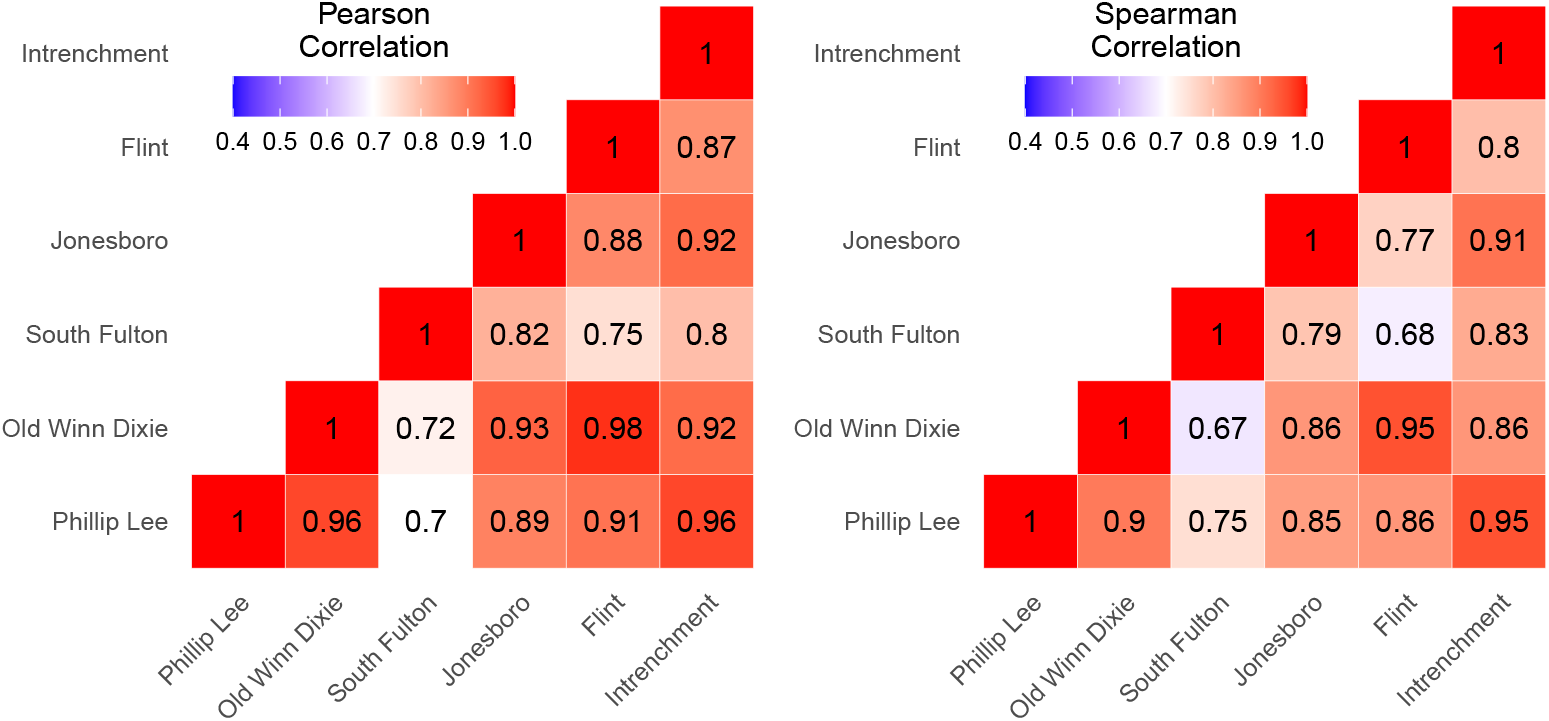
Correlations between SARS-CoV-2 RNA concentrations in the wastewater samples from six influent line sites.

**Supplementary Figure 3.**
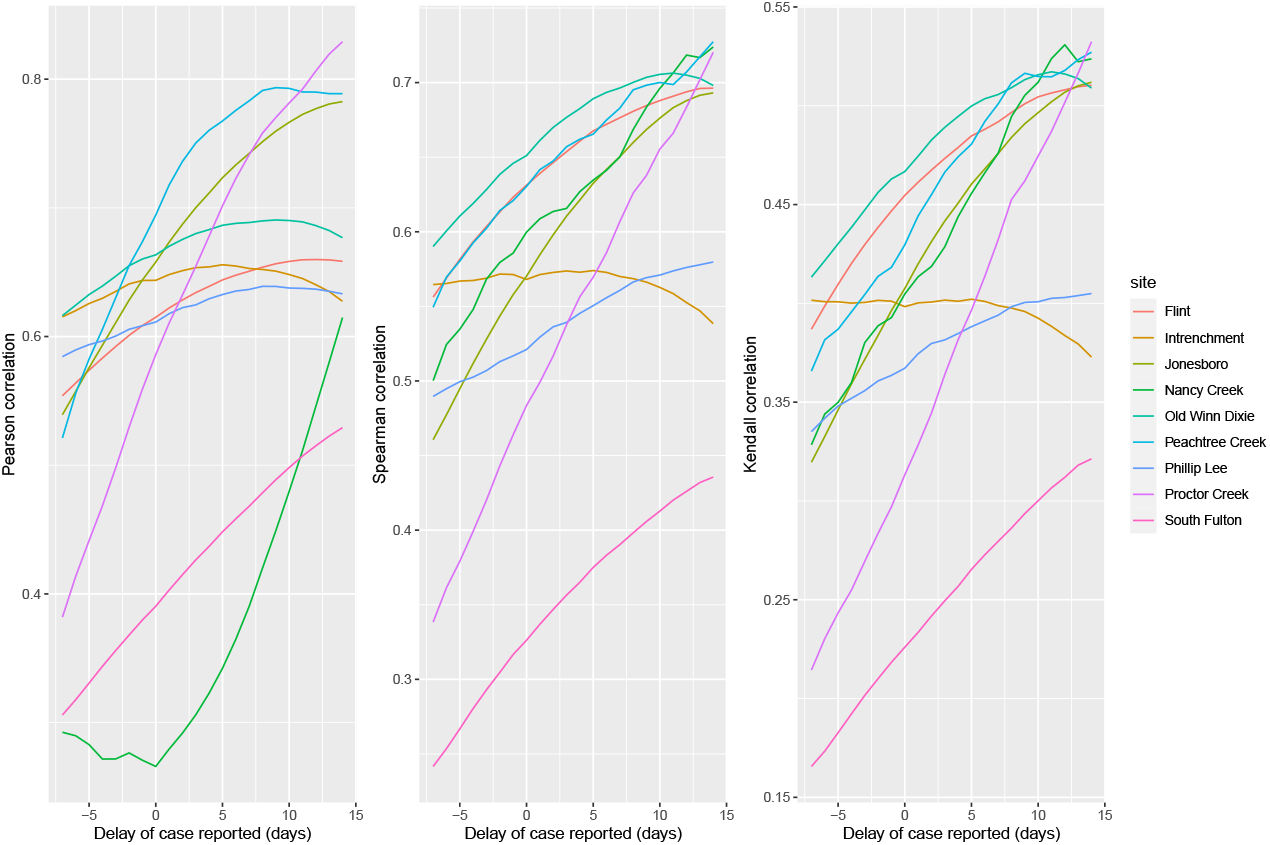
Correlations between SARS-CoV-2 RNA concentrations in the wastewater samples and delayed reported case numbers in Fulton County by influent line site.

**Supplementary Figure 4.**
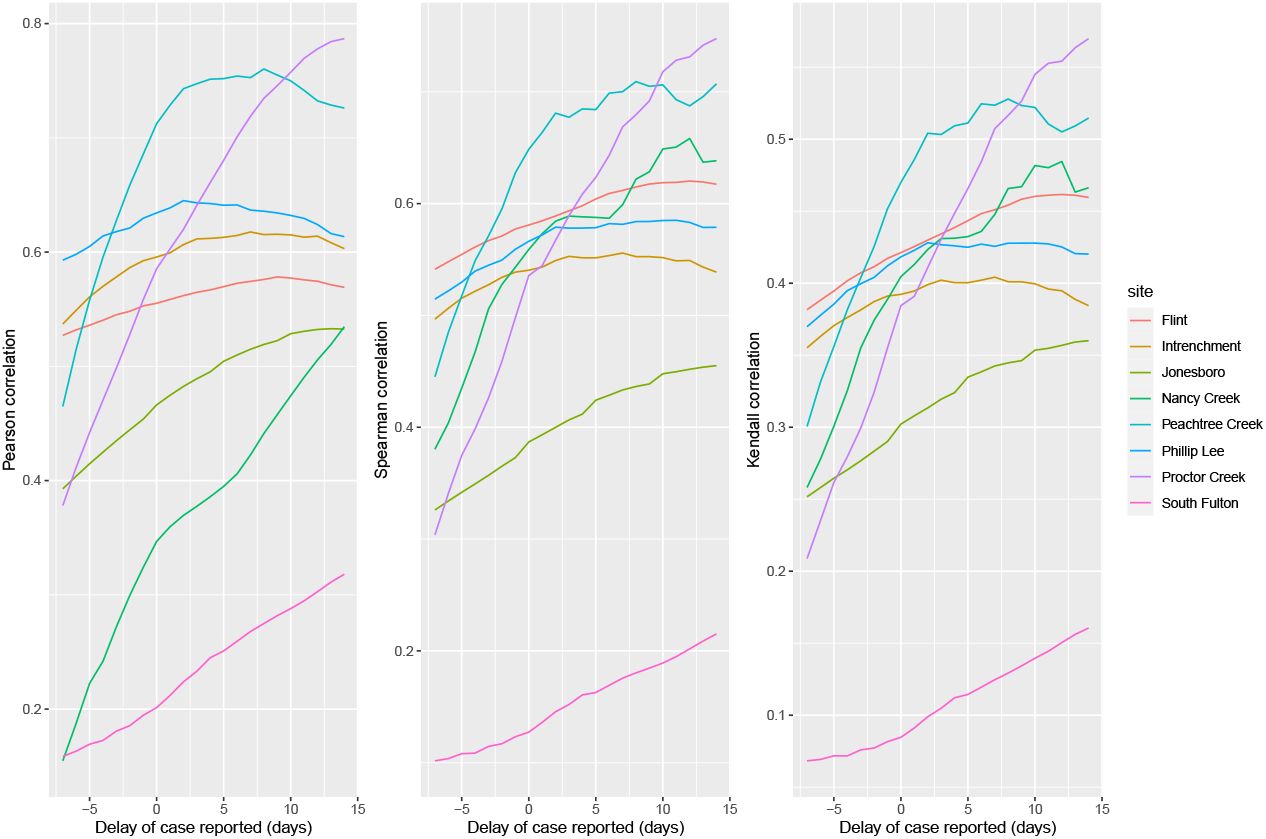
Correlations between SARS-CoV-2 RNA concentrations in the wastewater samples and delayed reported case numbers in the influent line catchment area by influent line site.

**Supplementary Figure 5.**
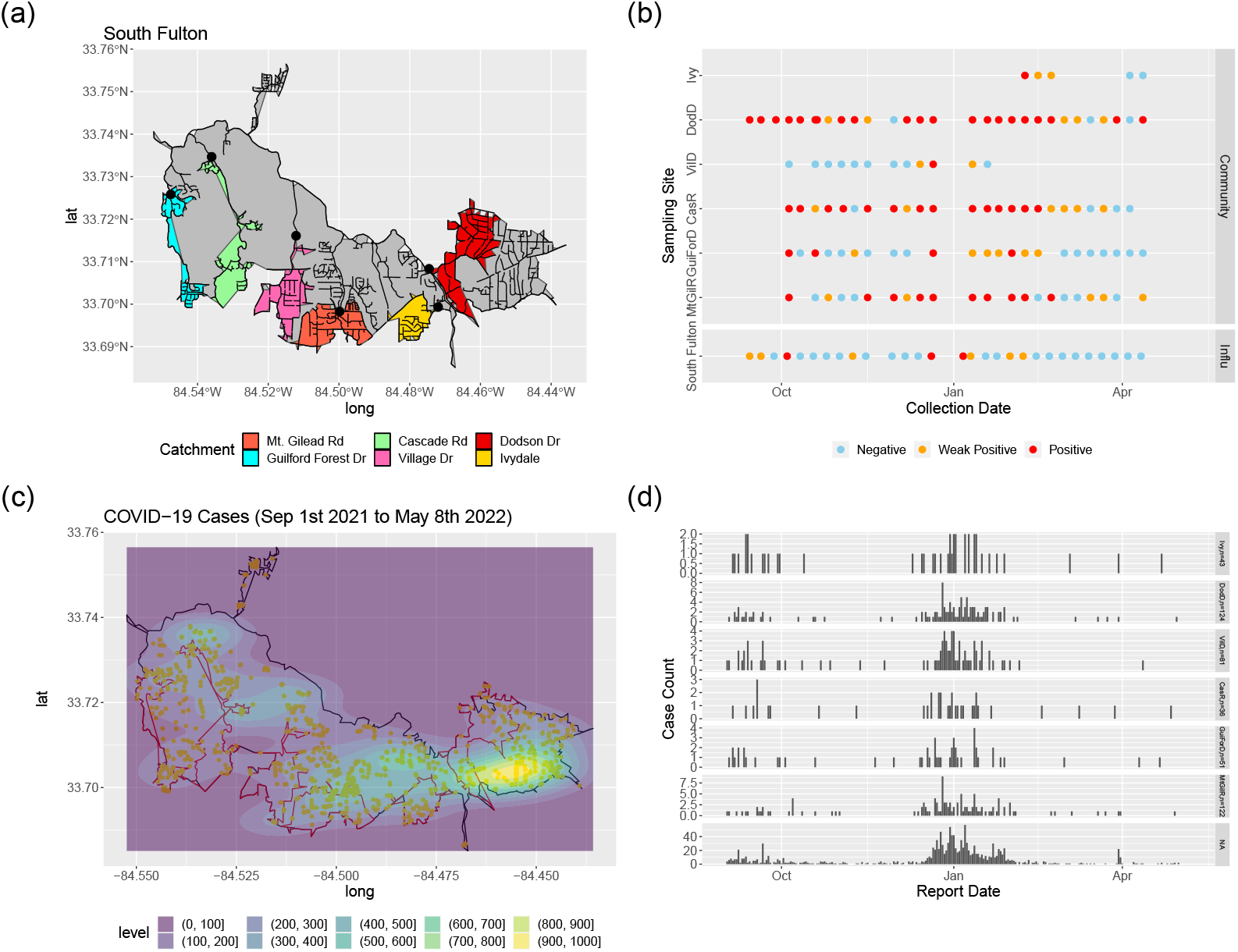
SARS-CoV-2 wastewater surveillance results for community sites and reported COVID-19 case numbers in the catchment area for South Fulton sampling cluster between September 2021–May 2022. Subfigure (a) shows the catchment areas of each community site nested within the overall catchment of the influent line site (in gray). The black lines represent the sewer network lines. Subfigure (b) shows the weekly wastewater surveillance results (RT-PCR detection of SARS-CoV-2 RNA). Subfigure (c) shows the heatmap of reported COVID-19 cases between September 1st 2021–May 8th 2022 within the influent catchment area. Subfigure (d) shows the epidemic curves of COVID-19 within each community site catchment area and NA represents all the cases in the catchment area of the influent line site that are not in the catchment area of a specific community site. MtGilR, GuiForD, CasR, VilD, DodD, and Ivy represent Mt. Gilead Rd, Guilford Forest Dr, Cascade Rd, Village Dr, Dodson Dr, and Ivydale respectively.

**Supplementary Figure 6.**
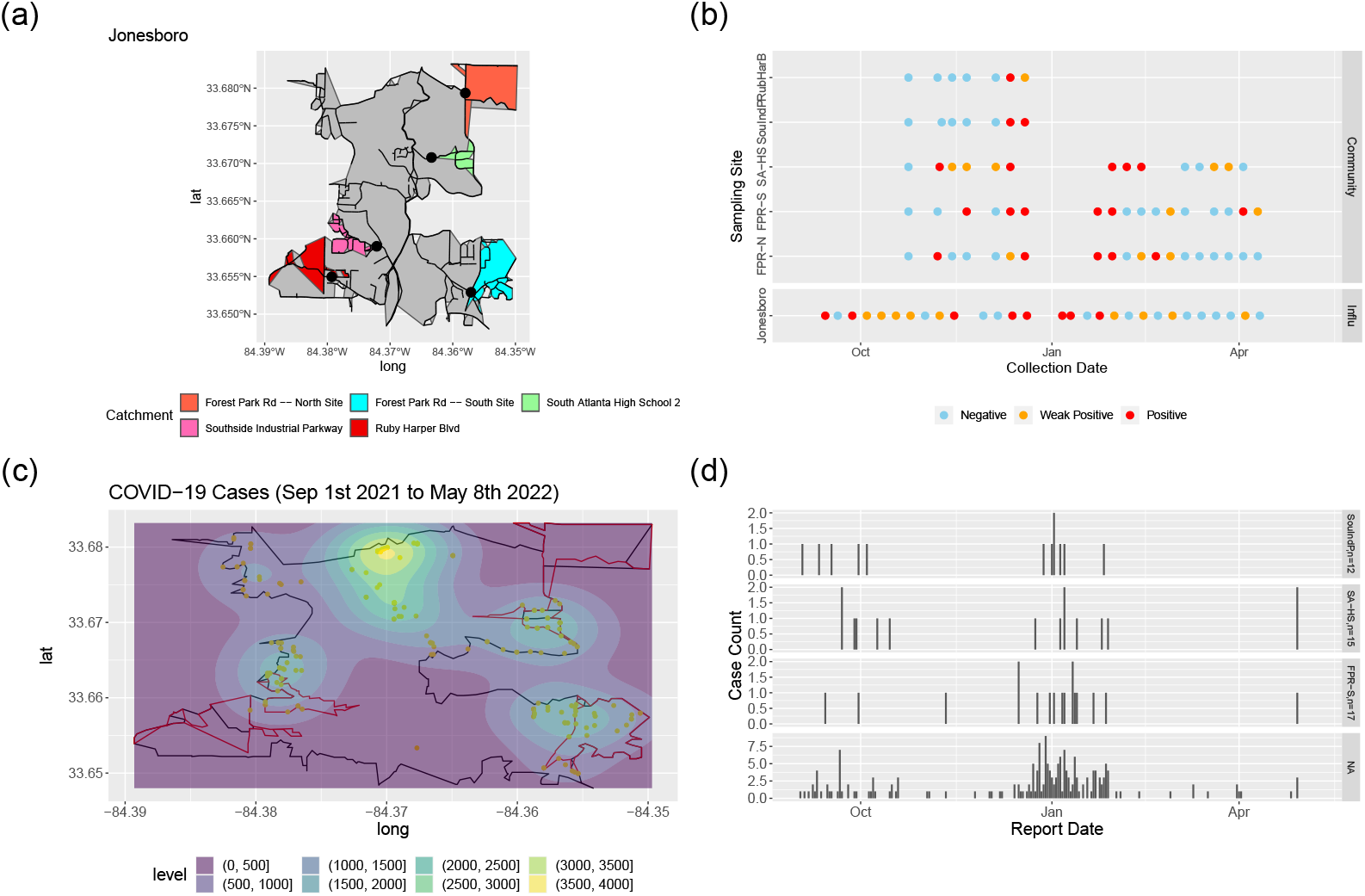
SARS-CoV-2 wastewater surveillance results for community sites and reported COVID-19 case numbers in the catchment area for Jonesboro sampling cluster between September 2021–May 2022. Subfigure (a) shows the catchment areas of each community site nested within the overall catchment of the influent line site (in gray). The black lines represent the sewer network lines. Subfigure (b) shows the weekly wastewater surveillance results (RT-PCR detection of SARS-CoV-2 RNA). Subfigure (c) shows the heatmap of reported COVID-19 cases between September 1st 2021–May 8th 2022 within the influent catchment area. Subfigure (d) shows the epidemic curves of COVID-19 within each community site catchment area and NA represents all the cases in the catchment area of the influent line site that are not in the catchment area of a specific community site. FPR-N, FPR-S, SA-HS, SouIndP, and RubHarB represent Forest Park Rd – North Site, Forest Park Rd – South Site, South Atlanta High School, Southside Industrial Parkway, and Ruby Harper Blvd respectively.

**Supplementary Figure 7.**
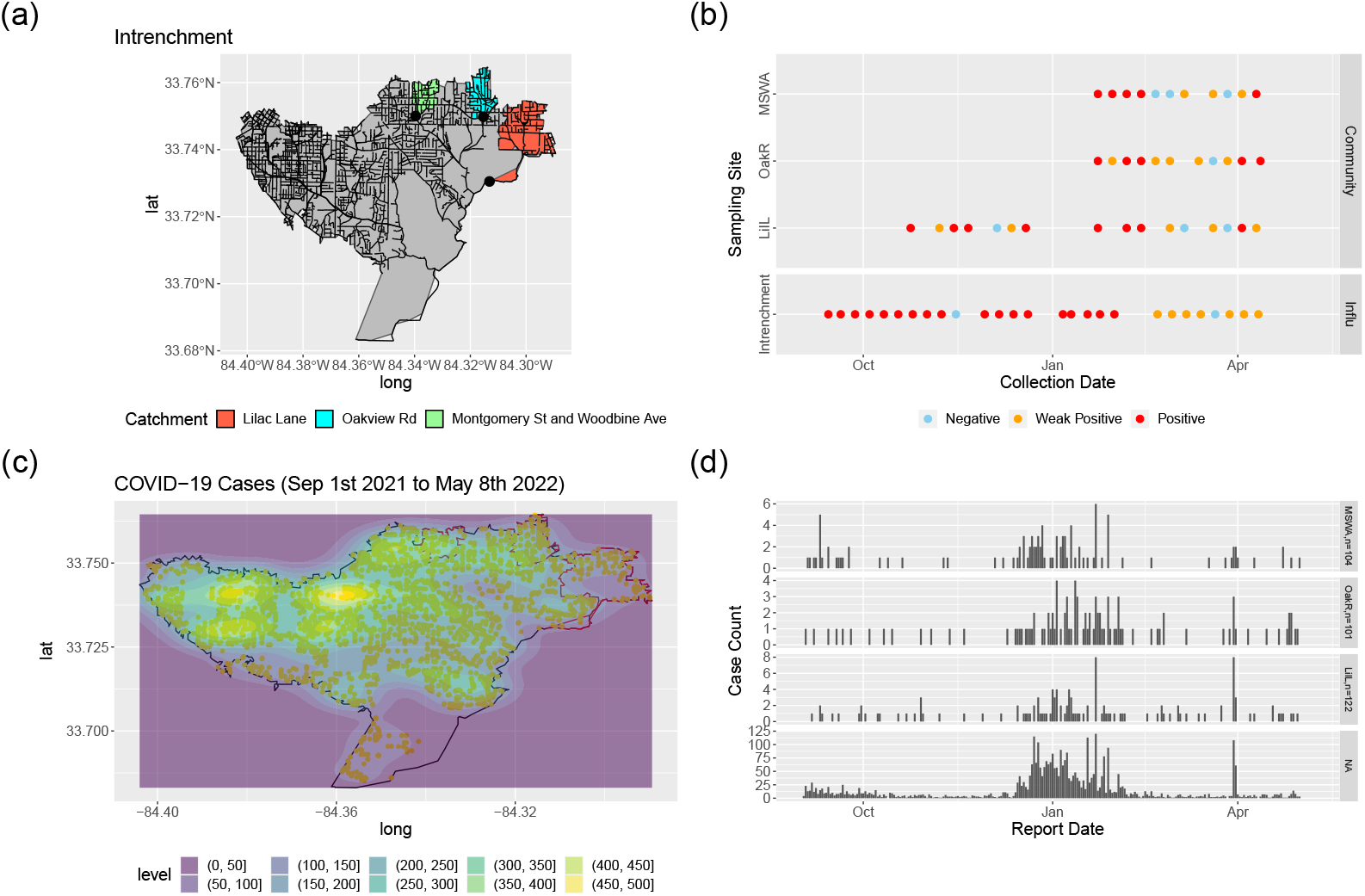
SARS-CoV-2 wastewater surveillance results for community sites and reported COVID-19 case numbers in the catchment area for Intrenchment sampling cluster between September 2021–May 2022. Subfigure (a) shows the catchment areas of each community site nested within the overall catchment of the influent line site (in gray). The black lines represent the sewer network lines. Subfigure (b) shows the weekly wastewater surveillance results (RT-PCR detection of SARS-CoV-2 RNA). Subfigure (c) shows the heatmap of reported COVID-19 cases between September 1st 2021–May 8th 2022 within the influent catchment area. Subfigure (d) shows the epidemic curves of COVID-19 within each community site catchment area and NA represents all the cases in the catchment area of the influent line site that are not in the catchment area of a specific community site. LilL, OakR, and MSWA represent Lilac Lane, Oakview Rd, and Montgomery St and Woodbine Ave respectively.

**Supplementary Table 1.**
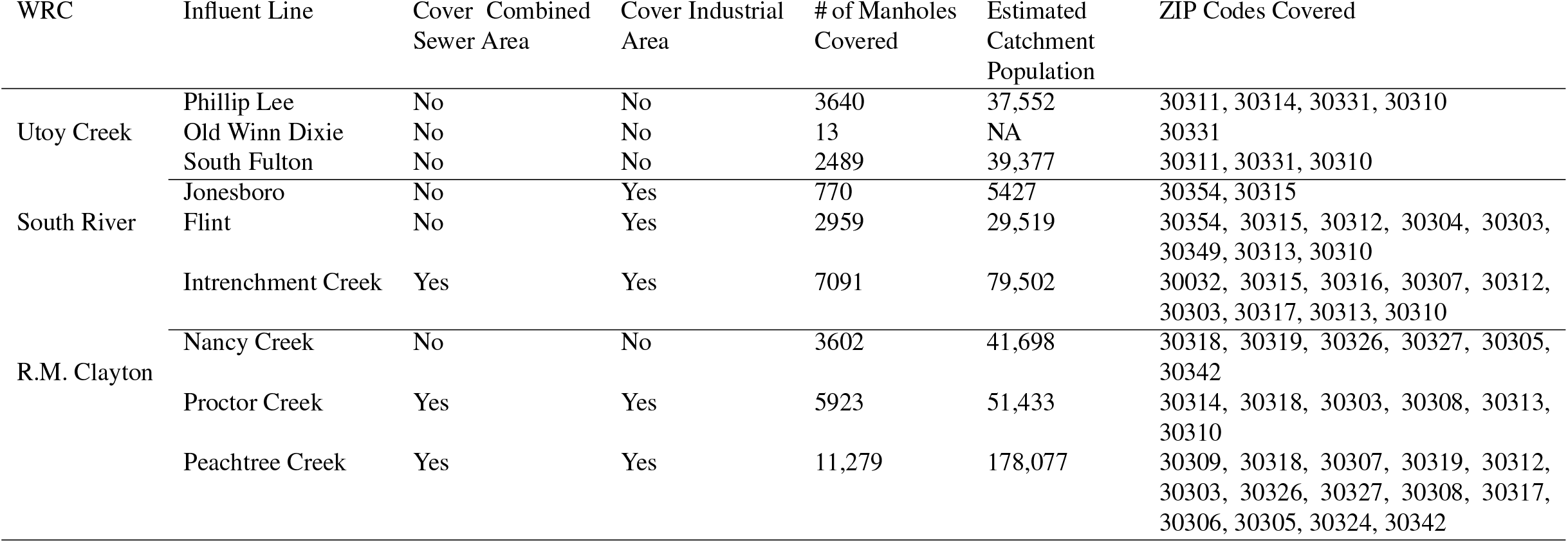
Descriptive information for influent lines. WRC represents Water Reclamation Center.

**Supplementary Table 2.**
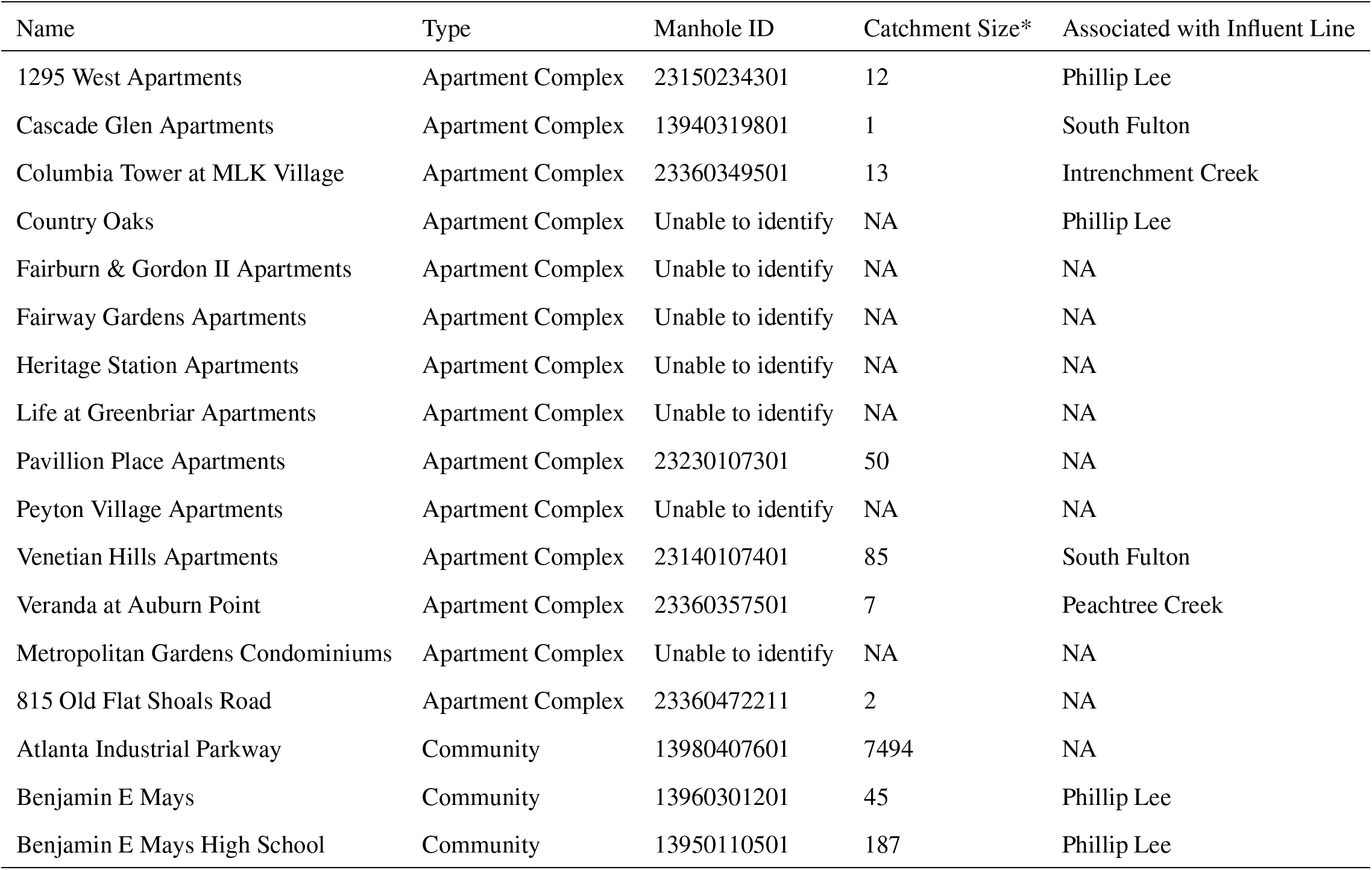
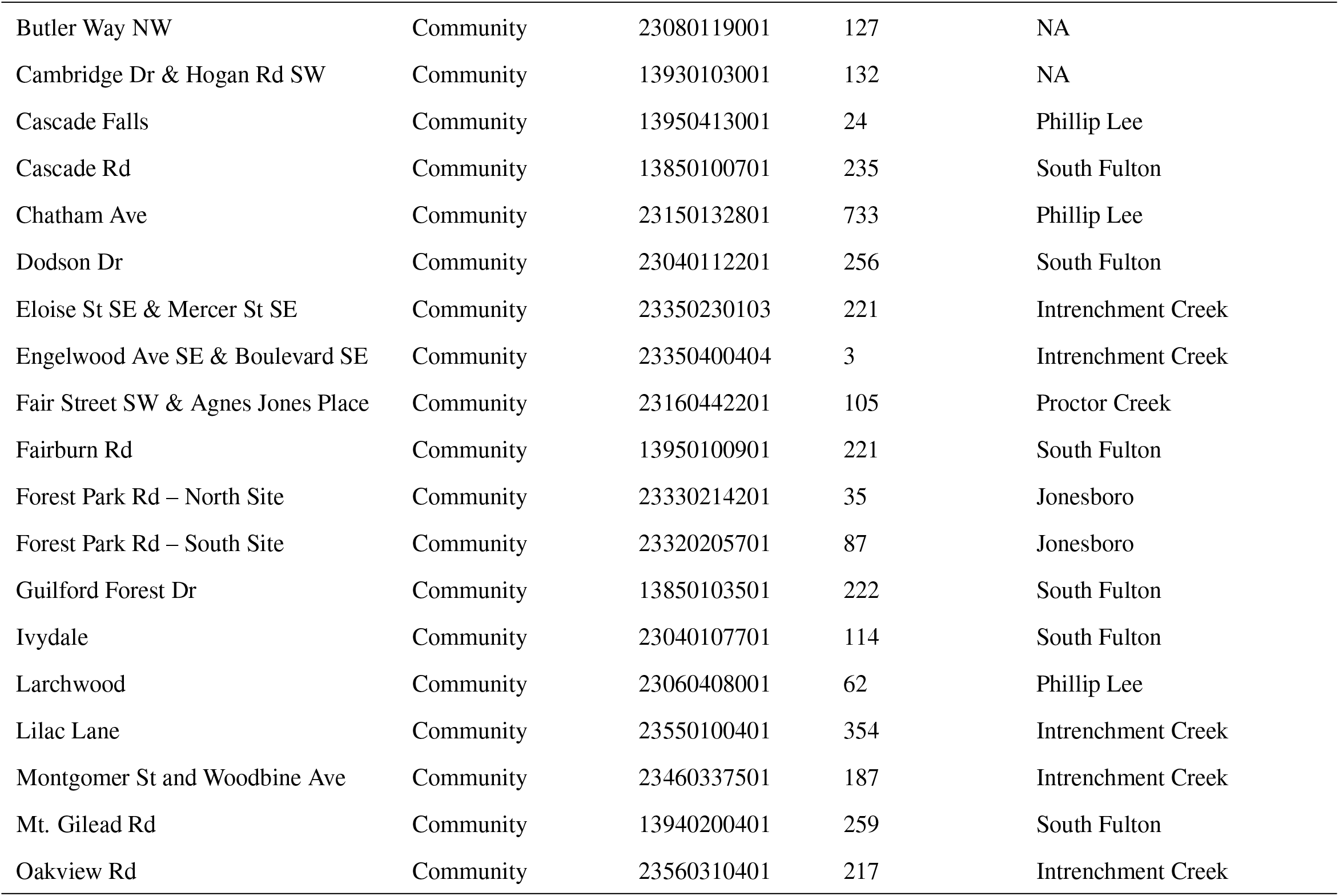
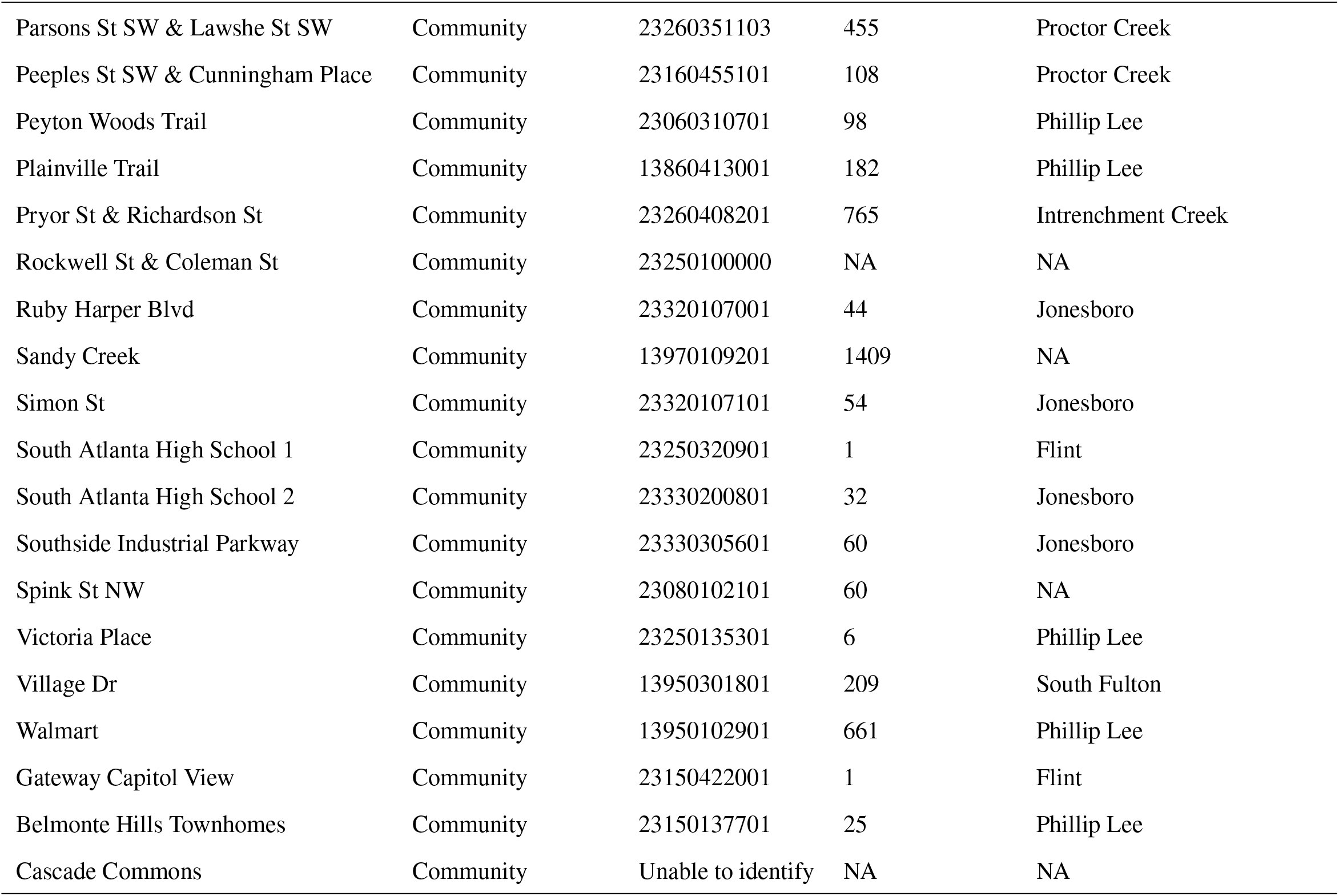
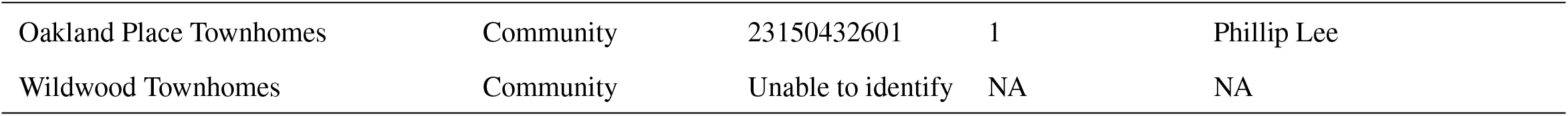
Descriptive information for community sites. *The catchment size are calculated as the number of manholes covered.

